# Genetic Analysis and Natural History of Parkinson’s Disease Due to the LRRK2 G2019S Variant

**DOI:** 10.1101/2023.10.26.23297636

**Authors:** Matthew J. Kmiecik, Steven Micheletti, Daniella Coker, Karl Heilbron, Jingchunzi Shi, Keaton Stagaman, Teresa Filshtein Sonmez, Pierre Fontanillas, Suyash Shringarpure, Madeleine Schloetter, Helen M. Rowbotham, Paul Cannon, Janie F. Shelton, David A. Hinds, Joyce Y. Tung, 23andMe Research Team, Michael V. Holmes, Stella Aslibekyan, Lucy Norcliffe-Kaufmann

## Abstract

The *LRRK2 G2019S* variant is the most common cause of monogenic Parkinson’s Disease (PD); however, questions remain regarding the penetrance, clinical phenotype, and natural history of carriers.

We performed a 3.5 year prospective longitudinal online study in a large number of 1,286 genotyped *LRRK2 G2019S* carriers and 109,154 controls, with and without Parkinson’s disease (PD) recruited from the 23andMe Research Cohort. We collected self-reported motor and non-motor symptoms every six months, as well as demographics, family histories, and environmental risk factors. Incident cases of PD (phenoconverters) were identified at follow-up. We determined lifetime risk of PD using accelerated failure time modeling and explored the impact of polygenic risk on penetrance. We also computed the genetic ancestry of all *LRRK2 G2019S* carriers in the 23andMe database and identified regions of the world where carrier frequencies are highest.

We observed that despite a one year longer disease duration (*p*=0.016), *LRRK2 G2019S* carriers with PD had similar burden of motor symptoms, yet significantly fewer non-motor symptoms including cognitive difficulties, REM sleep behavior disorder (RBD), and hyposmia (all *p-values*<0.0002). The cumulative incidence of PD in *G2019S* carriers by age 60 was 8.66%. *G2019S* carriers had a 10-fold risk of developing PD versus non-carriers. This rose to a 27-fold risk in *G2019S* carriers with a PD polygenic risk score in the top 25% versus non-carriers in the bottom 25%. In addition to identifying ancient founding events in people of North African and Ashkenazi descent, our genetic ancestry analyses infer that the *G2019S* variant was later introduced to Spanish colonial territories in the Americas.

Our results suggest *LRRK2 G2019S* PD appears to be a slowly progressive predominantly motor subtype of PD with a lower prevalence of hyposmia, RBD, and cognitive impairment. This suggests that the current prodromal criteria, which are based on idiopathic PD, may lack sensitivity to detect the early phases of *LRRK2* PD in *G2019S* carriers. We show that polygenic burden may contribute to the development of PD in the *LRRK2 G2019S* carrier population. Collectively, the results should help support screening programs and candidate enrichment strategies for upcoming trials of *LRRK2* inhibitors in early-stage disease.

## Introduction

Gain of function variants in the leucine-rich repeat kinase 2 gene (*LRRK2, PARK8*) are associated with monogenic Parkinson’s Disease (PD). The *LRRK2 G2019S* founder mutation is the most common cause of monogenic PD, with a prevalence estimate in the United states of up to 2.1%^1^, an autosomal dominant pattern of inheritance, and high but incomplete penetrance.^2^ The variant is most prevalent in Ashkenazi Jews and North African Berbers, where it is assumed that multiple founding events contributed to the high carrier rate in the two ancestral populations.^3,4^ The LRRK2 cytoplasmic protein has a large and complex structure that functions as both a kinase and GTPase.^5^ The *G2019S* missense variant, although located within the kinase domain,^2,6^ is thought to influence both enzymatic activities.^7^ Increased LRRK2 kinase activity has also been observed in postmortem brain tissue in idiopathic PD,^8^ leading to the hypothesis that LRRK2 inhibition might present a promising therapeutic strategy for PD in general.

The natural history of PD in *LRRK2* carriers is not well understood due to small sample sizes and the lack of compatible prospective data sets. The lifetime prevalence of PD in *LRRK2 G2019S* carriers is hotly debated, with penetrance estimates ranging from 25% to 80%.^1,9^ Precise phenoconversion rates have not been established.^10^ Validating sensitive and specific prodromal criteria to detect the early phases of *LRRK2* PD, as the symptoms of neurodegeneration are emerging, will be important to design enriched neuroprotective trials in at-risk carriers.^11,12^ As it is not known which combination of symptoms are the first to emerge, it is unclear if the prodromal criteria derived from idiopathic PD apply to patients with *LRRK2* PD,^13^ as these weigh heavily both rapid eye movement sleep behavior disorder (RBD) and hyposmia as high risk markers.^14,15^ Yet, small cohort studies in *LRRK2 G2019S* carriers with PD suggest that RBD is less prevalent^16^ and reports of anosmia are mixed, with some *LRRK2* PD patients having preserved olfaction.^13,17^

In addition, the clinical course of *LRRK2* PD may progress differently. Observations suggest that *LRRK2* PD is a slowly progressive predominantly motor subtype of PD^13,18^ with low rates of dementia,^19^ and better survival.^20^ While patients with idiopathic PD that develop dementia have the hallmark misfolded α-synuclein-containing intraneuronal Lewy body inclusions throughout the limbic system and neocortex at autopsy,^21^ at least one-third of *LRRK2 G2019S* carriers with PD do not exhibit this classic Lewy body pathology anywhere in the brain, including in the substantia nigra.^19^ More recent studies show that unlike cases of idiopathic PD, *LRRK2* carriers are more likely to have a negative cerebral spinal fluid α-synuclein seeding amplification assay test, in which the α-synuclein from *LRRK2* patients does not misfold and promote fibrillization.^12^ Taken together, these studies indicate a difference in the core pathophysiology of *LRRK2* PD, where at least in a subset of patients their neuronal injury appears to be independent of α-synuclein.

The 23andMe, Inc. research participant database is the largest pre-existing genetic cohort developed via direct-to-consumer genetic testing that contains data on human disease, and uniquely poised to understand *LRRK2* PD. In 2018, we began an initiative to enroll genotyped *LRRK2 G2019S* carriers into a prospective online natural history study. We compared phenotypic and genotypic differences between carriers and non-carriers with and without PD. We used genetic ancestry to map where the founder mutation arose and understand the migratory patterns. We prospectively identified incident cases and used survival analyses to estimate cumulative incidence of PD among carriers and non-carriers. In addition, we investigated the impact of polygenic risk scores (PRS) on susceptibility to PD in the context of *LRRK2 G2019S*. Currently, these results provide the largest prospective natural history study of *LRRK2 G2019S* PD that spans geographical ancestry, phenotypic, and genotypic features.

## Materials and Methods

### Participants

23andMe’s Parkinson’s Impact Project (PIP) was launched in 2018 with the goal of conducting an online survey-based longitudinal prospective study of *LRRK2 G2019S* carriers. Recruitment emails explaining the purpose of the study were sent to all eligible *LRRK2 G2019S* carriers from 23andMe Research cohort. All study research participants were >18 years old, US residents, and provided informed consent to volunteer to participate (protocol approval: AAHRPP-accredited Salus IRB). Specific eligibility criteria for the *LRRK2 G2019S* participants included: 1) confirmation of at least one *LRRK2 G2019S* allele; 2) having opted into receiving and opening their 23andMe FDA-approved *LRRK2 G2019S* carrier status report (that included a referral for genetic counseling); and 3) permission to be recontacted about research opportunities. Recruitment into the cohort continued on a rolling basis. Eligible non-carriers were randomly selected from the 23andMe Research cohort. Participants included in the analysis were enrolled between November 2018 until October 4^th^, 2022.

### Study Design

Surveys were administered through the 23andMe website and/or mobile application. The baseline survey surfaced to carriers and non-carriers captured: (i) demographics (age, sex, education), (ii) PD diagnosis and family history, (iii) motor symptoms, (iv) mood/depression/cognition, (v) sleep, (vi) autonomic symptoms, and (vii) lifestyle/environmental exposures. To capture symptom progression, *LRRK2 G2019S* carriers were recontacted every 6 months for 3.5 years (i.e., seven follow-up surveys, see Fig. 1). Non-carriers were recontacted every 12 months to complete a Health Update survey and report any new PD diagnosis. Phenoconverters (i.e., incident cases) were defined as carrier and non-carrier participants that reported “no” to a PD diagnosis at baseline (study entry), but subsequently reported “yes” to a new PD diagnosis at any subsequent encounter (>7 days after baseline completion).

**Fig. 1.**
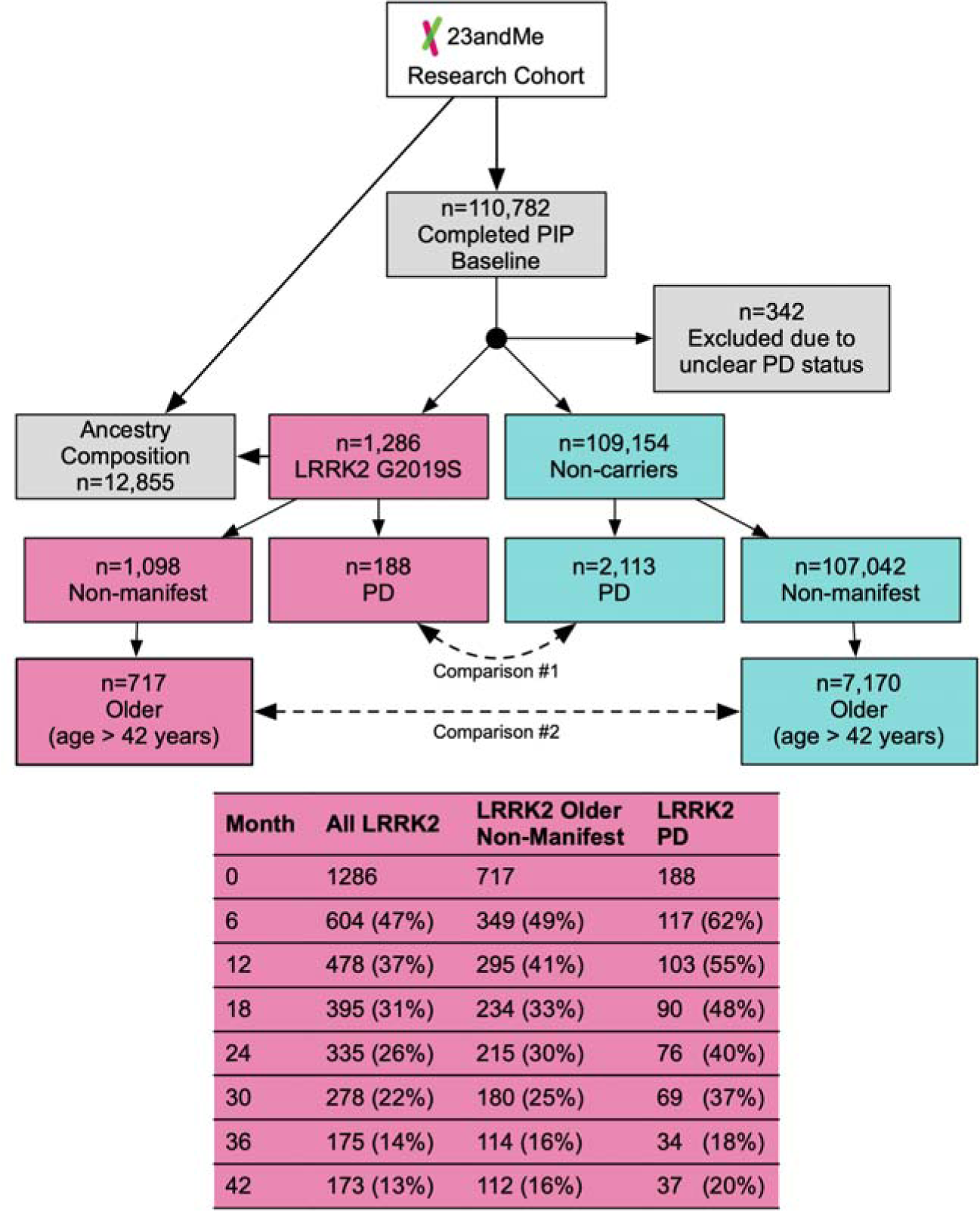
Parkinson’s Impact Project study design and analysis plan. Flow chart that shows recruitment of consented participants, classification according to *LRRK2 G2019S* carrier status and PD diagnosis. The older non-manifest subgroup was selected based on age at risk of PD (>42 years, i.e., within 2*SD* of age of PD diagnosis for *LRRK2 G2019S* carriers). *LRRK2 G2019S* carriers received a similar survey every 6 months. Non-carriers were asked to report any new diagnosis of PD every 12 months. Sample sizes and percentage rates of follow-up survey completions are shown for the entire *LRRK2 G2019S* cohort, older non-manifest *LRRK2 G2019S* carriers, and *LRRK2 G2019S* carriers with PD. Note the higher rates of follow-up in *LRRK2 G2019S* PD and older non-manifest *LRRK2 G2019S* carriers. Pink=*LRRK2 G2019S* carriers; Blue=non-carriers.

### Measures

#### Genetic Data

DNA extraction and genotyping were performed on saliva samples by Clinical Laboratory Improvement Amendments-certified and College of American Pathologists-accredited clinical laboratories of Laboratory Corporation of America. Samples were genotyped on one of five genotyping platforms (see the Supplementary Materials for details). The accuracy of genotyping the *LRRK2 G2019S* and *GBA N370S* variants was 99% [97-100%] with >99% reproducibility and repeatability.^22^ *LRRK2 G2019S* carrier status was established by the presence of the genotyped G2019S variant.

#### Motor and Cognitive Symptoms

Motor deficits were assessed by asking participants to self-report having tremor, imbalance/falls, gait changes (bradykinesia, shuffling, festination; freezing of gait; dragging feet, and reduced arm swing), stooped posture; hypophonia; and deterioration in handwriting. Cognition and executive functioning were assessed with self-reported difficulties with attention/concentrating, distractibility, multitasking, completing tasks, or understanding over the last 12 months. Diagnosis of mild cognitive impairment (MCI) was collected. Memory was assessed by self-reported worsening of forgetfulness, word recall, remembering the date, or misplacing things over the last 12 months. Depression was assessed using a modified version of the PHQ-9.^23^

#### Prodromal Markers

We followed the Movement Disorders Society Research Criteria for signs and symptoms of prodromal PD, using proxy answers when needed.^15^ The presence of dream re-enactment behavior was assessed by asking participants whether they reported acting out dreams or had previously been diagnosed with RBD. Olfaction was assessed by asking participants to rate their current ability to smell, with those that reported “very poor” or “poor” smell being classified as having possible hyposmia. Autonomic symptoms were assessed over the last 3 months and included orthostatic hypotension (lightheadedness), constipation (<3 bowel movements/week), urinary dysfunction (urgency or frequency) and erectile dysfunction. Daytime somnolence was derived from the PHQ-9 in participants that reported more than half of the time trouble falling/staying asleep, sleeping too much, feeling tired or having little energy.

#### Risk Markers

Six questions were administered to assess lifetime environmental exposures to pesticides/solvents/heavy metals in the workplace/home, and employment requiring the use of a respirator, mask or gloves. Occupational toxin exposure was classified as responding “yes” to regular exposure to pesticides, including herbicides, fungicides, insecticides, rodenticides, or fumigants at work. Occupational PPE (personal protective equipment) use was classified as responding “yes” to ever having regularly used a respirator, mask, or gloves at the workplace. Participants that did not report weekly consumption of coffee were classified as low-caffeine intake. Following the established standardized definition,^24^ non-smoking status was determined as smoking <100 cigarettes in a lifetime. Physical inactivity was determined as participants that reported they did not participate weekly in any form of physical activity. Family history of PD was defined as having a first-degree relative with PD. Lifetime history of head injury with loss of consciousness due to sports, accidents or violence was also captured.

### Data Analysis

#### Baseline Comparisons

Participants were categorized according to their *LRRK2 G2019S* status (i.e., carrier vs. non-carrier) and PD diagnosis (i.e., manifest vs. non-manifest) creating four comparison groups: 1) *LRRK2 G2019S* PD (manifest carriers), 2) *LRRK2 G2019S* non-manifest (*LRRK2 G2019S* carriers without PD), 3) non-carrier PD (manifest PD without the variant), and 4) non-carrier controls (non-carriers without PD). To account for age as a risk factor for PD, we created two subgroups of non-manifest participants (see Fig. 1). First, we selected a subgroup of older non-manifest *LRRK2 G2019S* carriers that had an age of at least 42 years. This lower-bound age cutoff was determined by calculating 2*SD* below the mean age of PD diagnosis for carriers. Next, we performed 10:1 age/sex matching using a nearest-neighbor propensity score algorithm^25^ to select a comparison group of older non-carrier controls matched to the older non-manifest *LRRK2 G2019S* carriers (see Fig. 1). The main statistical comparisons were: 1) *LRRK2 G2019S* PD vs. non-carrier PD and 2) older *LRRK2 G2019S* non-manifest carriers vs. older non-carrier controls. Within each comparison, we performed logistic regressions to model the effects of *LRRK2 G2019S* status. P-values were corrected for multiple comparisons using false discovery rate^26^ (FDR; L=.05) within each category (i.e., demographics, motor symptoms, prodromal markers, risk factors). Using the same matching procedure, we created a third group of non-carrier controls age- and sex-matched to all PD cases (i.e., both *LRRK2 G2019S* carriers and non-carriers) for descriptive purposes.

#### Ancestry Composition

To understand the ancestral origins and genetic structure of *LRRK2 G2019S* carriers, we performed a suite of ancestry analyses on all research participants who carry the *G2019S* variant. We determined the frequency of carrier prevalence around the world by comparing all research participants with *LRRK2 G2019S* in the 23andMe database (*n*=12,855) to research participants who reported their grandparent birth locations (*n*=545,187 outside of the US; *n*=2,162,527 inside the US). Because a subset of research participants reported grandparent birth locations at sub-national levels, we also performed kernel density estimation to determine sub-nation geographic regions that have high densities of carriers. We identified segments of DNA that are identical by descent (IBD) between all individuals using templated Positional Burrows–Wheeler Transform (TPBWT) IBD detection.^27^ TPBWT is robust to genotype and phasing errors and its default parameters are optimized to maximize accuracy on the custom designed Illumina BeadChip genotyping array. Consequently, we ran TPBWT with default parameters, but only retained IBD segments sizes larger than 5cM to minimize potential false-positive estimates.^27^ We identified genetic clusters of *LRRK2 G2019S* carriers using Leiden community detection on the total amount of shared IBD between individuals. We first pruned close relatives in the dataset (individuals that share ≥ 700cM of IBD) to avoid producing clusters that correspond to close familial relationships. The Leiden algorithm determines clusters of individuals who share a recent common ancestor by maximizing modularity, or within-group IBD sharing related to between-group IBD sharing.^28^ We labeled any identified genetic clusters using enrichment of geographic birth locations of members’ grandparents. Finally, we employed 23andMe’s Ancestry Composition algorithm to infer local ancestry in genomic windows across each individual’s chromosome. Ancestry Composition predicts the genomic origin of 45 local populations that are nested within broader geographic categories and is optimized for high coverage genotype array data.^29^

#### Neuroanatomical Modeling of PD

Survey items were grouped across six domains (cognitive, memory, autonomic, motor, smell, and RBD) and differences in symptom prevalence were compared in *LRRK2 G2019S* PD vs. non-carrier PD using logistic regressions accounting for disease duration, age, sex, and education as covariates. Logistic regressions without covariates were separately computed using weights from coarsened exact matching^30^ to confirm results. FDR correction was applied to each model term. Further, we built an anatomical-based model of neuropathology by associating each of the six symptom domains with structural areas of the brain using the Braak framework^31^ to create a spatial representation of pathological deficits. According to the frequency of neurological symptoms reported, we color coded according to aggregated symptom burden in the substantia nigra (motor), cortex (cognition/memory), subarea of the pons (RBD),^32^ brainstem/periphery (autonomic), and olfactory bulb (hyposmia) to visually model patterns of neurodegeneration and recapitulate the neurobiology of PD.

#### Disease-free Survival Analysis

We used Kaplan-Meier estimation to describe time to PD diagnosis survival curves (i.e., using self-reported age of PD diagnosis) between *LRRK2 G2019S* carriers and non-carriers. To include prodromal risk factors, we estimated an accelerated failure time (AFT) model with a Weibull hazard shape. In addition to *LRRK2 G2019S* status, we included the following known PD risk factors as binary variables: occupational toxin exposure, male sex, caffeine non-user, non-smoker, previous head injury with loss of consciousness. We included education (≥ Associate degree) as a binary covariate. Participants without a PD diagnosis after 42 months or those lost to follow-up were right censored and age at their last completed survey was used as the time of censoring. Phenoconversion rates were estimated in participants who were over 40 years old at the time of study entry, as this age group is considered to be at risk.

#### Polygenic Risk Score Analysis

We computed a polygenic risk score (PRS) for each participant using allelic weights from the most recently published GWAS meta-analysis of PD (*n*=37.6k PD cases, *n*=1.4 million controls, *n*=18.6k proxy-cases).^33^ 23andMe contributed GWAS summary statistics for *n*=2.4k cases (6.5% of the total cases in the GWAS) and *n*=571.4k controls (41%) to this meta-analysis.^34^ The overlap between 23andMe research participants used in the recent GWAS used to derive the PRS and the PIP cohort was small: 13% *LRRK2 G2019S* non-manifest, 9% *LRRK2 G2019S* PD, 6% non-carrier PD, and 7% non-carrier controls; hence, overlapping participants were not removed from the present analyses.

To synthesize a PRS that could be evaluated in both carriers and non-carriers of *LRRK2* variants, we took the original PRS (1,839 variants) and removed SNPs ±10 Mb surrounding the *LRRK2* gene, which included 46 proximal variants, to compute a modified PRS (see the Supplementary Materials for details). Both the original and modified PRS were scaled to have a mean of 0 and a standard deviation of 1 (i.e., Z-score) using the entire PIP cohort.

We computed two logistic regressions (i.e., one for each PRS) separately for *LRRK2 G2019S* carriers and non-carriers predicting PD status as a function of PRS with baseline age, sex, and 10 principal components (PCs) of ancestry as covariates (see Supplementary Materials for details). We visually inspected the PCs and determined that the first 10 PCs demonstrated separation and structure for use as covariates in logistic regressions (see Supplementary Fig. 1).

To examine the relationship between the PRS and risk of PD, we excluded participants under 40 years of age and split the *LRRK2 G2019S* carriers and non-carriers into six groups depending on the modified PRS percentile ranges: low (1-25%), intermediate (25-75%), and high (75-100%; see Fahed et al.^35^). We used logistic regression to model PD status as a function of PRS group with baseline age, sex, and ancestry PCs as covariates. Non-carriers with intermediate PRS value served as the reference group. To examine the impact of polygenic versus monogenic risk in greater detail, we explored the association with PD risk across deciles of the modified PRS, among *LRRK2 G2019S* carriers and non-carriers, adjusting for baseline age, sex, and ancestry PCs. Predicted odds ratios across carrier status were estimated using non-carriers with median (fifth decile) PRS as a reference group and mean values of the remaining covariates. To test deviations from additivity, we compared deciles models with and without the *LRRK2 G2019S* carrier status by PRS interaction term using analysis of variance. PRS analyses used both baseline and incident cases of PD. Given that the Nalls et al.^33^ allelic weights were derived from a European-centric GWAS, all PRS analyses were repeated excluding non-Europeans to compare in our heterogeneous, but majority European, sample. The European-only analyses were identical except that only five ancestry PCs were used as covariates in addition to age and sex (see Supplementary Fig. 2).

#### Software

Data analysis and visualization was performed using R^36^ (v. 3.6). Case-control matching was performed using *MatchIt*^25^ and verified with *cem.*^30^ Survival analyses were performed using *survival*^37^ and *flexsurv.*^38^ Figures were prepared using *ggplot2.*^39^

## Results

### Baseline Characteristics

Table 1 summarizes the participant characteristics at study entry (see Supplementary Table 1 for additional groups). Fig. 1 describes the participant flow and sample sizes of the comparison groups. Relative to non-carriers, the *LRRK2 G2019S* carrier cohort (*n=*1,286) were more likely to be female, have a higher level of education, be of Ashkenazi Jewish descent, and have a first degree relative with PD. *LRRK2 G2019S* carriers were also more likely to carry the *GBA N370S* variant.

**Table 1.**
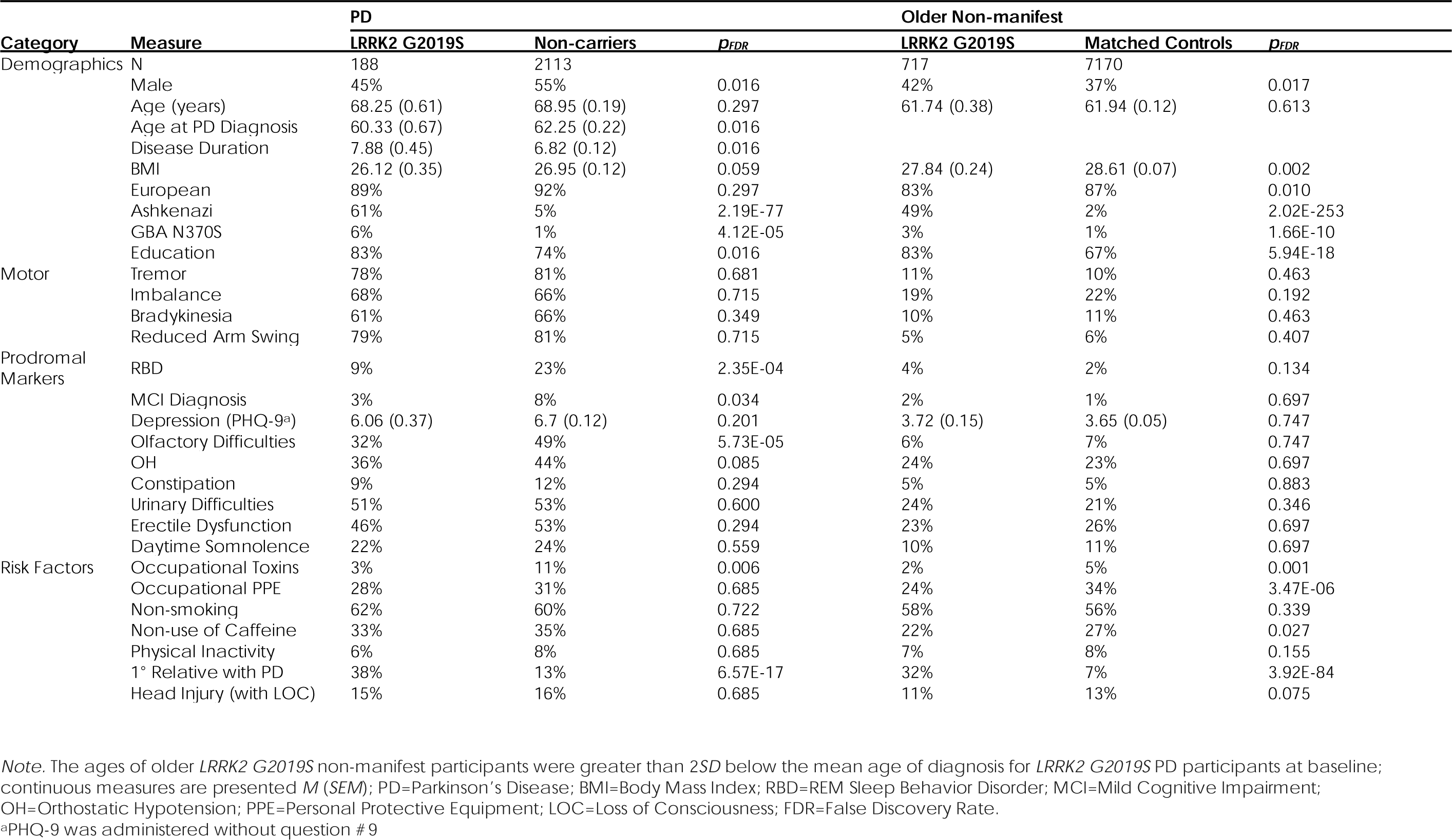
Clinical Characteristics of the Parkinson’s Impact Project Cohort at Study Entry.

#### Ancestry of LRRK2

Geographical analyses in Fig. 2A showed that most *LRRK2 G2019S* carriers had grandparents born around the southern coastal regions of the Mediterranean basin, with the strongest signals in Maghreb, including Algeria (2.5% prevalence), Morocco (2.1%), and Tunisia (1.7%); eastern Europe, including modern day Ukraine (0.7%) and Belarus (1.2%); Latin America, including Puerto Rico (0.5%), Cuba (0.6%), Colombia (0.3%), and Mexico (0.1%) (see Supplementary Table 2). Leiden community detection on shared identical by descent (IBD) segments identified seven genetic groups in G2019S carriers (modularity = 0.525) that correspond to geography and genetic ancestry (see Supplementary Table 3). These groups included Ashkenazim from Eastern Europe (*n*=6,918), North Africa (*n*=399), Italy (*n*=265), Northwestern Europe (*n*=1,926), Puerto Rico (*n*=695), Cuba (*n*=877), and Mexico (*n*=646). The remaining 1,129 individuals were assigned to clusters with sample sizes ≤ 20 individuals and did not contain enough metadata to label. By arranging genetic groups by the mean amount of IBD shared, Ashkenazim from Eastern Europe were most genetically similar to North Africa (see Fig. 2B). Puerto Rico, Mexico, and Cuba groups clustered together but were most genetically similar to Eastern Europe, suggesting founder effects from migrations out of Europe into the Americas. Northwestern Europe and Italy were the most distantly connected, suggesting the mutation entered these regions through more distant migrations. Within genetic groups, ancestry inference indicated a higher proportion of Ashkenazi genetic ancestry in the Eastern Europe group, North African genetic ancestry in the North Africa group, and Iberian genetic ancestry in the Latin American groups (see Fig. 2C). However, North African and Ashkenazi ancestry was present across individuals within the Latin American groups (see Supplementary Fig. 3).

**Fig. 2.**
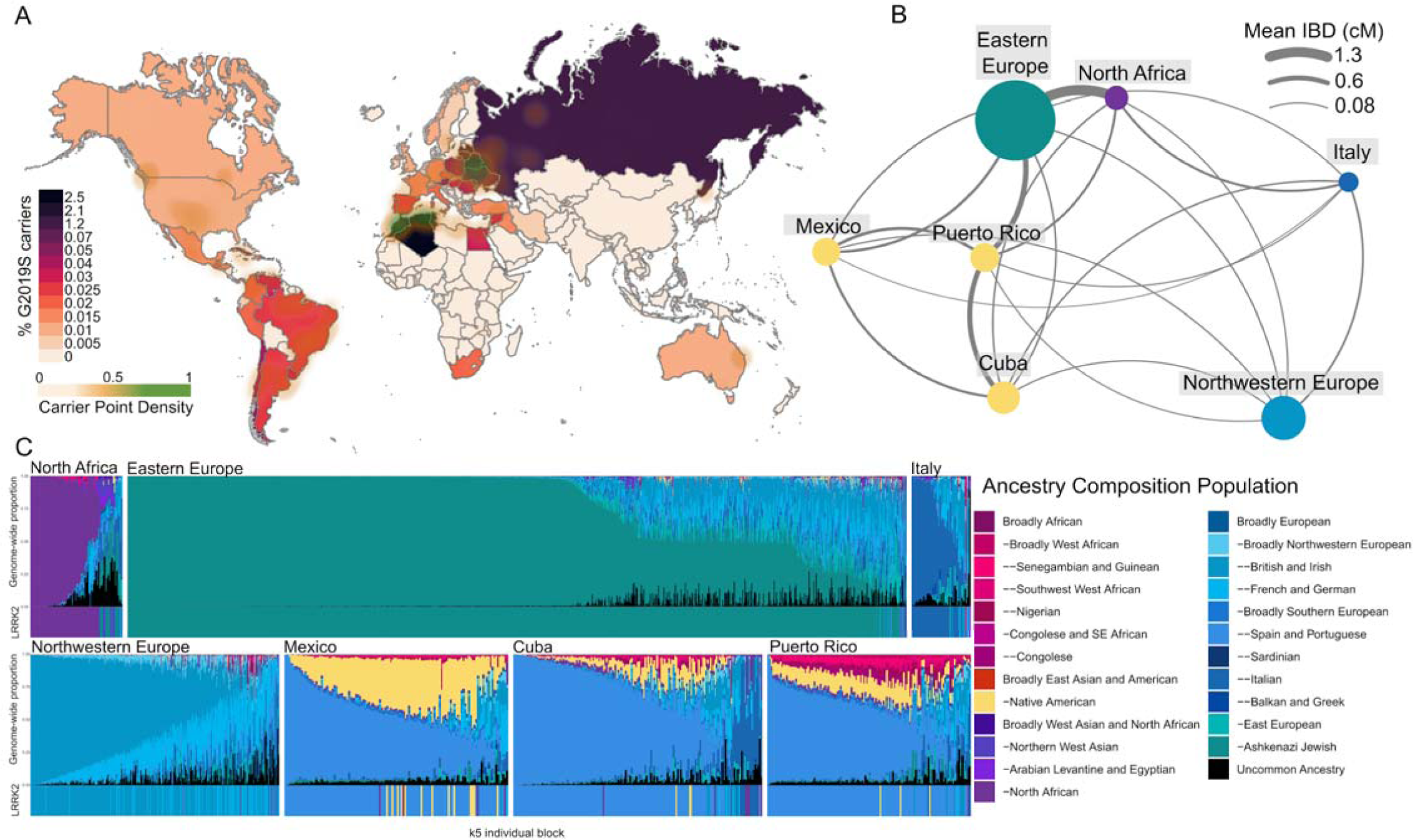
Ancestry of *LRRK2 G2019S* carriers. Ancestry and geographic origins of individuals who carry the *LRRK2 G2019S* mutation. A) Frequency of *LRRK2 G2019S* carriers based on participant-reported grandparent birth countries. Green clouds correspond to sub-national locations with the highest frequency of carriers based on kernel density estimates. B) The genetic structure of the seven groups identified in *LRRK2 G2019S* carriers. Groups are arranged in a graph using ForceAtlas2 orientation based on the mean pairwise identical by descent (IBD) sharing rate. C) Ancestry Composition of carriers belonging to each genetic group. The mean genome-wide ancestry of individuals is displayed above, with the ancestry at *LRRK2* reflected below.

#### PD: LRRK2 G2019S Carriers vs. Non-carriers

At entry, *n*=188 *LRRK2 G2019S* carriers and *n*=2,113 non-carriers reported a diagnosis of PD (see Fig. 1). Lifestyle risk factors were similar between the groups (i.e., non-smoking, non-use of caffeine, head injuries, and physical inactivity). Despite being of similar age at study entry, *LRRK2 G2019S* carriers reported their age of PD diagnosis ∼2 years younger (*p*=0.027) than non-carriers, resulting in a longer disease duration at the time of entry. Fig. 3 shows differences between symptom prevalence in *LRRK2 G2019S* PD and non-carrier PD (see Supplementary Table 4). *LRRK2 G2019S* PD participants had a similar burden of motor symptoms to PD non-carriers yet reported significantly fewer non-motor symptoms. Compared to non-carrier PD, *LRRK2 G2019S* PD reported lower rates of RBD (23% vs. 9%), lower rates of olfactory deficits (49% vs. 32%), and fewer mild cognitive impairment (MCI) diagnoses (8% vs. 3%). No differences in autonomic symptoms were observed. Modeling of symptom domains with underlying anatomical mapping showed that the motor regions were equally impacted, but suggested less pathology outside the substantia nigra in *LRRK2 G2019S* PD compared to non-carrier PD.

**Fig. 3.**
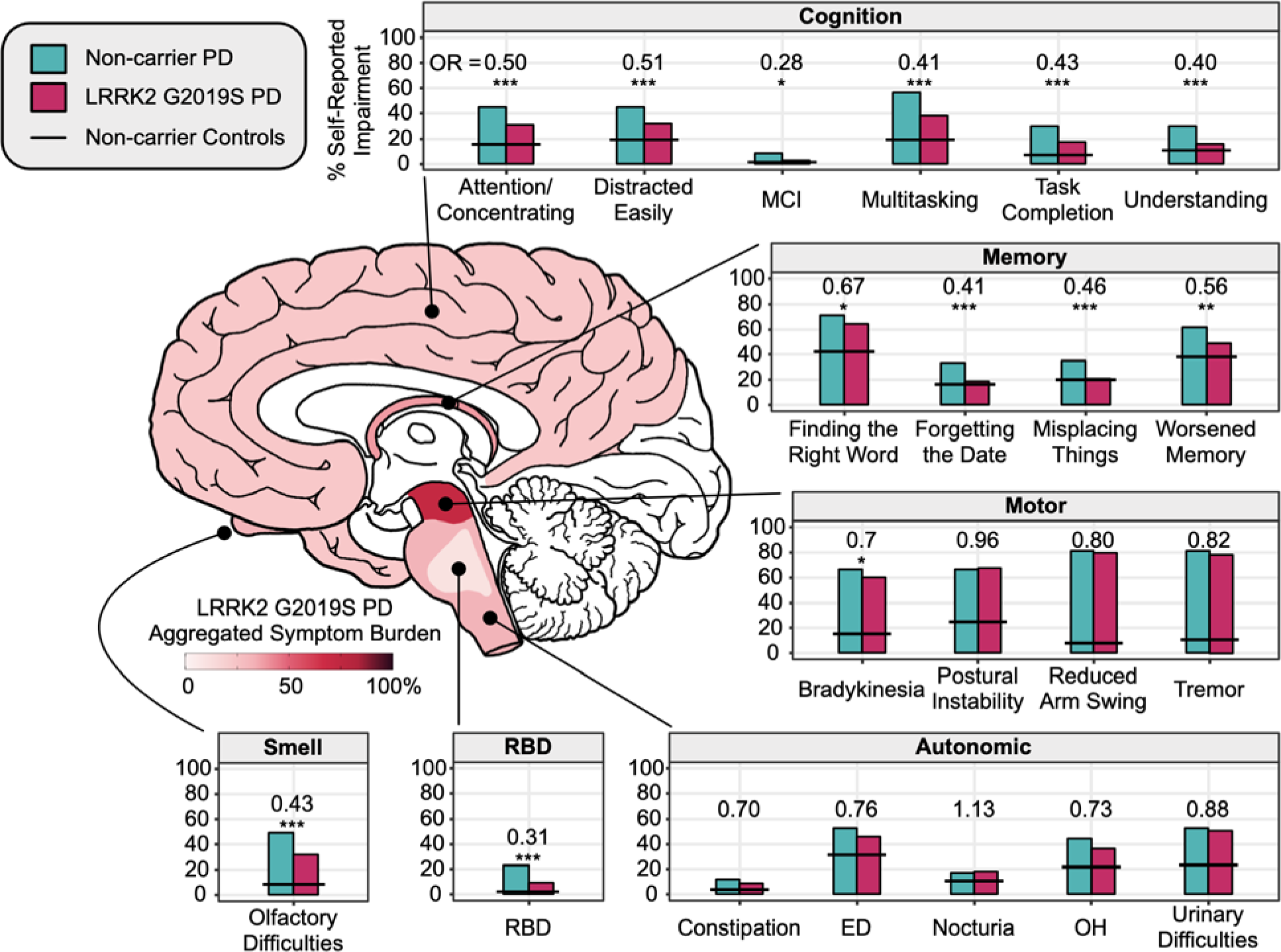
Relative to non-carrier PD, *LRRK2 G2019S* PD reported lower symptomatic burden in cognitive, memory, autonomic, RBD, and olfactory domains. Symptoms were clustered by domain. Bars show the percentage of *LRRK2 G2019S* PD carriers and non-carrier PD that self-reported a symptom. Differences between groups were estimated using odds ratios from logistic regressions adjusted for age, sex, education, and disease duration. Horizontal black lines show prevalence reported by age- and sex-matched non-carrier controls (not included in modeling). Schematic of the brain shows corresponding neuropathological sites underlying symptom burden for *LRRK2 G2019S* PD. Aggregated symptom burden was calculated by averaging the percentage of reported symptoms across each phenotype domain in *LRRK2 G2019S* PD. Note the similarities in the reported frequency of motor symptoms, but lower prevalence for symptoms corresponding to regions outside the basal ganglia (e.g., RBD, smell, cognition). \**p*<0.05; \*\**p*<0.01; \*\*\**p*<0.001; OR=odds ratio comparing *LRRK2 G2019S* PD and non-carrier PD; RBD=REM sleep behavior disorder; MCI=mild cognitive impairment diagnosis; ED=erectile dysfunction; OH=orthostatic hypotension; PD=Parkinson’s Disease.

### Phenoconversions to PD

In participants without a PD diagnosis at entry, there were no differences in the rates of reported symptoms in older *LRRK2 G2019S* carriers (*n*=717) and matched older non-carrier controls (*n*=7,170). Over the 3.5 years of follow-up in asymptomatic participants >40 years of age, we observed five newly diagnosed cases of PD in *LRRK2 G2019S* carriers and 53 new cases in non-carriers. Thus, prospectively *LRRK2 G2019S* carriers had an estimated 10-times the risk of developing PD compared to non-carrier controls (0.19 vs. 0.019%/year). Put differently, we observed five incident cases of PD per 1000 person-years in *LRRK2 G2019S* carriers. *LRRK2 G2019S* carriers that developed PD were on average 6.6 years (*SEM*=2.0 years) younger in age at PD diagnosis compared to non-carriers newly diagnosed with PD. Prior to diagnosis, the most commonly reported non-motor symptoms reported by the non-carriers that subsequently developed PD included RBD diagnosis/dream reenactment behavior (13%) and smell loss (27%). In contrast, none of the five *LRRK2 G2019S* cases reported suspected RBD prior to their PD diagnosis. Prodromal autonomic symptoms reported in non-carrier prior to diagnosis included erectile dysfunction in men (48%), urinary difficulties (43%), and orthostatic hypotension (37%). There was no clear pattern of prodromal autonomic symptoms in *LRRK2 G2019S* carriers prior to phenoconversion.

### PD-free Survival

Fig. 4 shows PD-free survival curves with Kaplan-Meier estimation of the cohort stratified according to their *LRRK2 G2019S* carrier status. We computed a Weibull AFT model that included known prodromal risk factors of PD on a subset of participants who had non-missing data across all covariates: *n*=772 *LRRK2 G2019S* non-manifest, 162 *LRRK2 G2019S* PD, 1,574 non-carrier PD, and 78,445 non-carrier controls. The AFT model showed that *LRRK2 G2019S* carrier status had the greatest impact on the age of PD diagnosis (see Table 2). The estimated coefficient for carriers was −0.32 95% CI [−0.35, −0.30], suggesting an accelerated rate of receiving a PD diagnosis. Correspondingly, the time ratio of 0.72 indicates that, on average, *LRRK2 G2019S* carriers were diagnosed with PD approximately 28% earlier compared to non-carriers. Other known PD risk factors had detectable, but comparably modest effects on PD-free survival rates (3-11%).

**Fig. 4.**
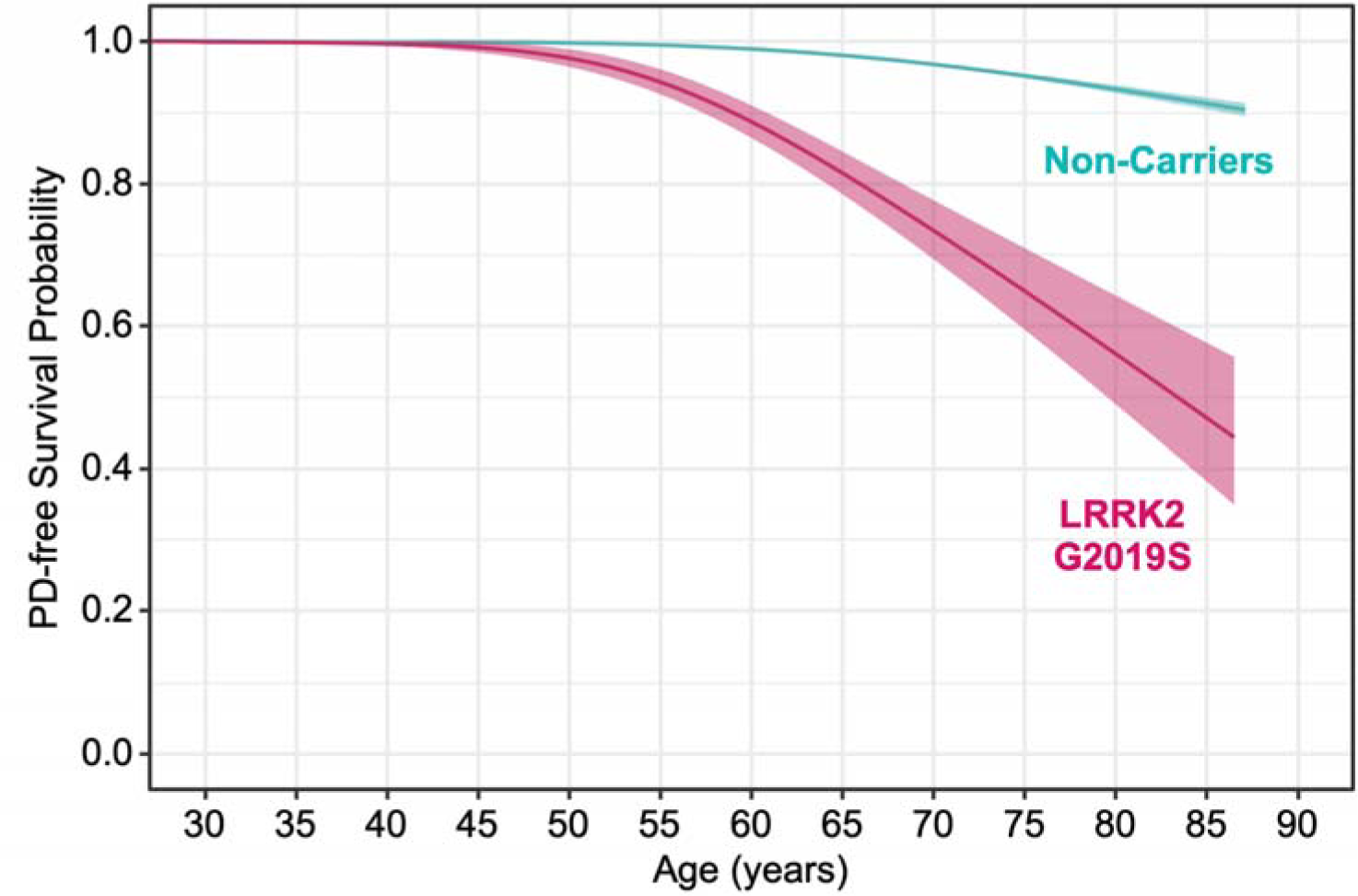
Survival curves with Kaplan-Meier estimation show a steeper decline in PD-free survival in *LRRK2 G2019S* carriers (*n*=1,154) relative to non-carriers (*n*=97,308). Curves were smoothed using generalized additive models to protect data privacy. Shading depicts 95% confidence intervals.

**Table 2.** Weibull Accelerated Failure Time Model Coefficients and Time Ratios of Parkinson’s Disease-free Survival.

| Term | Coefficient | 95% CI [LL UL] | SE | W | Time Ratio | 95% CI | p |
| --- | --- | --- | --- | --- | --- | --- | --- |
| LRRK2 G2019S | -0.32 | [-0.35 -0.3] | 0.013 | -24.72 | 0.72 | [0.71 0.74] | 7.02E-135 |
| Occupational Toxin Exposure | -0.11 | [-0.13 -0.09] | 0.012 | -9.61 | 0.89 | [0.87 0.91] | 7.30E-22 |
| Male | -0.06 | [-0.07 -0.05] | 0.007 | -8.30 | 0.94 | [0.93 0.96] | 1.02E-16 |
| Caffeine non-use | -0.06 | [-0.07 -0.04] | 0.007 | -7.76 | 0.94 | [0.93 0.96] | 8.76E-15 |
| Non-Smoking | -0.05 | [-0.07 -0.04] | 0.007 | -7.21 | 0.95 | [0.94 0.96] | 5.64E-13 |
| Previous Head Injury with LOC | -0.04 | [-0.05 -0.02] | 0.009 | -3.74 | 0.97 | [0.95 0.98] | 1.85E-04 |
| Education (≥ Associate Degree) | -0.03 | [-0.04 -0.01] | 0.008 | -3.22 | 0.97 | [0.96 0.99] | 1.26E-03 |
| Shape | 6.97 | [6.73 7.22] | 0.126 |  |  |  |  |
| Scale | 128.90 | [125.44 132.46] | 1.790 |  |  |  |  |

*Note.* CI=confidence interval; LL=lower level; UL=upper level; LOC=loss of consciousness.

### PRS

We identified similar odds ratios for associations between PD and both the original and modified PRS that excluded the *LRRK2* gene (see Supplementary Table 5; European-only results in Supplementary Table 6). To explore differences in polygenicity to PD risk among *LRRK2 G2019S* carriers and non-carriers, we compared the distributions adjusted for age, sex, and ancestry PCs of the modified PRS between individuals with and without PD. PD cases had a greater median PRS compared to those without a PD diagnosis for both carriers and non-carriers of *LRRK2 G2019S* (see Fig. 5A).

**Fig. 5.**
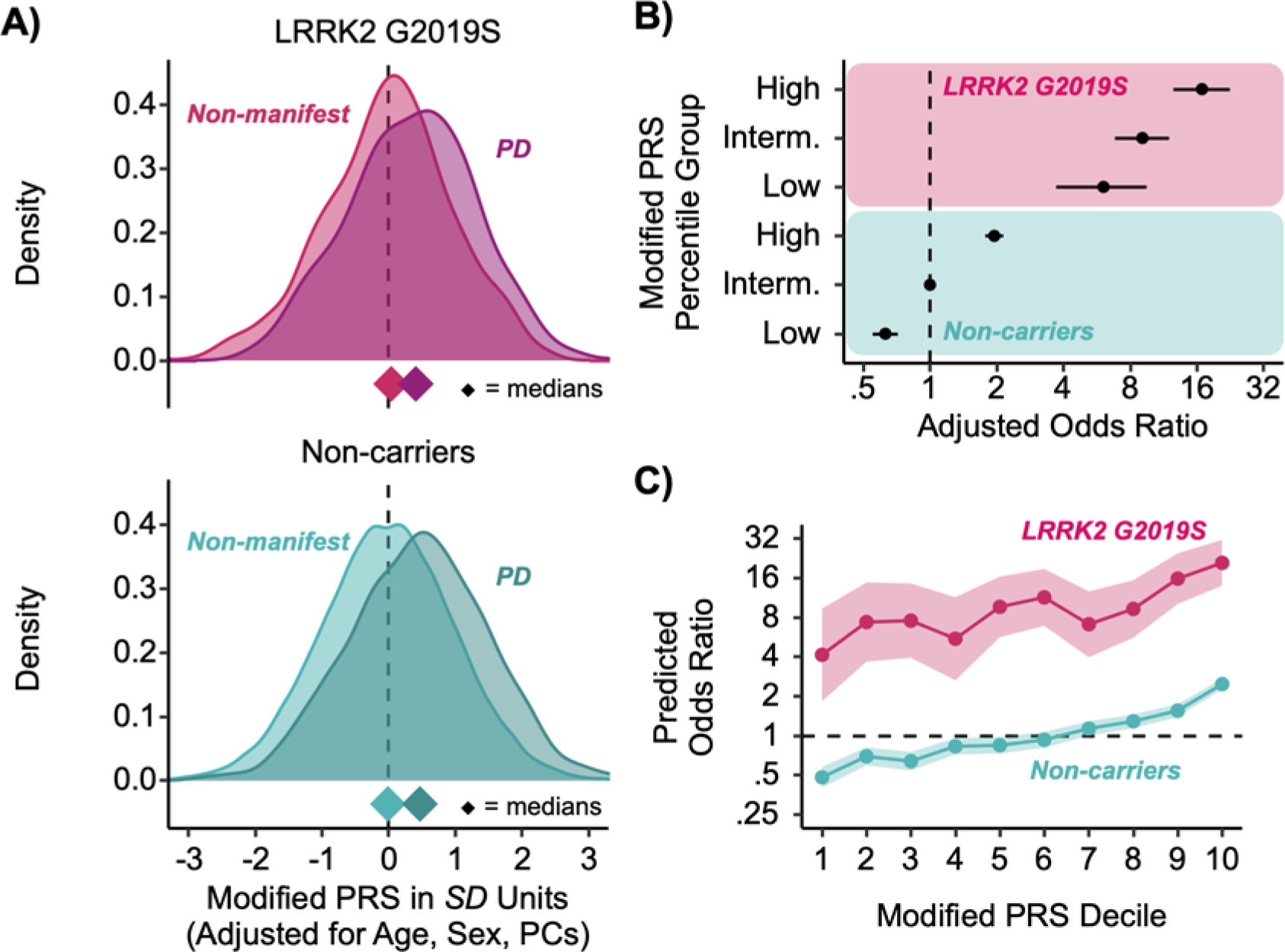
Associations between PRS and PD prevalence for participants > 40 years of age. A) Distributions of the modified PRS adjusted for age, sex, and ancestry PCs. Diamonds indicate median PRS. Non-manifest non-carriers were the non-carrier controls. B) Odds ratios of PD diagnosis between *LRRK2 G2019S* carriers and non-carriers stratified by PRS percentile groups: low (0-25%), intermediate (25-75%), high (75-100%). Logistic regressions were adjusted for age, sex, and ancestry PCs, and non-carriers with intermediate PRS were the reference group. Error bars are 95% confidence intervals (CIs). C). PD odds ratio between *LRRK2 G2019S* carriers and non-carriers referenced to non-carriers with median PRS across PRS deciles. Shading denotes 95% CIs. *Note*. PD=Parkinson’s Disease; PRS=polygenic risk score; Interm.=intermediate; PCs=ancestry principal components.

We examined dose-response relationships between the modified PRS and PD using logistic regression. Non-carriers at the median of the PRS served as the reference group. Both the group-wise (i.e., low, middle, high PRS) and decile models demonstrated a positive monotonic relationship between PRS and relative odds of PD (see Supplementary Table 7; see Supplementary Table 8 for European-only results). Compared to non-carriers in the middle of the PRS distribution (i.e., intermediate), non-carriers in the top 25% of the PRS had nearly double the relative odds of PD (*OR*=1.95 [1.78 2.14], *p*=2.03×10^-44^) and non-carriers in bottom 25% PRS had a lower relative odds of PD (*OR*=0.63 [0.55 0.72], *p*=4.08×10^-12^). Using the same reference group, the relative odds of PD among *LRRK2 G2019S* carriers was substantially greater than non-carriers. Compared to intermediate non-carriers, among *LRRK2 G2019S* carriers in the bottom 25% of the PRS, the relative odds was 6-fold greater (*OR*=6.06 [3.71 9.49], *p*=4.24×10^-14^), for those in the middle of the PRS it was 9-fold greater (*OR*=9.09 [6.82 11.99], *p*=2.94×10^-53^), and among those in the top 25% of the PRS it was 16-fold greater (*OR*=16.87 [12.55 22.55], *p*=7.73×10^-80^) (see Fig. 5B). Taking a more extreme comparison, *LRRK2 G2019S* carriers in the top 25% of the PRS had a 27-times greater relative odds of PD compared to non-carriers in the bottom 25% of the PRS.

Examining these associations in greater detail using deciles of the modified PRS relative to non-carriers at the fifth PRS decile (median) yielded predicted odds ratios that were between six-times (seventh decile) and 12x (sixth decile) greater across all PRS deciles among *LRRK2 G2019S* carriers compared to non-carriers (see Fig. 5C). For example, relative to non-carriers in the fifth PRS decile the predicted odds ratio among non-carriers in the top decile was 2.92 as compared to an *OR* of 24.69 among *LRRK2 G2019S* carriers in the same decile (see Supplementary Table 9; see Supplementary Table 10 for European-only results). An ANOVA comparing the additive model, which includes PRS decile and *LRRK2 G2019S* status as separate terms, to the interactive model, which includes their interaction, demonstrated that deviations from additivity were not observed in the decile model (*p*=0.71). Removing *GBA N370S* carriers did not change the findings. Taken together, these findings demonstrate the additive effect of LRRK2 on top of underlying polygenic risk of PD and highlights that among *LRRK2 G2019S* carriers polygenicity explains some of the variability in whether individuals develop PD. Similar results were obtained for participants with European-only ancestry.

## Discussion

In this study involving 1,286 *LRRK2 G2019S* carriers corresponding to 1,621-person-years of follow-up (1,292 person-years at risk of PD) and 109,154 non-carrier controls, we show that the predicted cumulative incidence of PD in *LRRK2 G2019S* carriers by the age of 60 is 8.66%. We show that the penetrance of the *LRRK2 G2019S* variant is influenced by polygenicity, with the risk of a PD diagnosis increasing 16-fold in carriers that also have a PRS within the top 25^th^ percentile relative to non-carriers in the middle range of the PRS (i.e., 25-75^th^ percentile). However, the *LRRK2 G2019S* variant still carries a risk above and beyond polygenicity, with carriers at lowest polygenic risk having greater relative odds of PD than non-carriers at the highest polygenic risk (see Fig. 5).

The clinical expression of *LRRK2 G2019S* PD appears to be different than in non-carriers with idiopathic PD. Overall, we found that *LRRK2 G2019S* carriers with PD were less likely to report non-motor features such as olfactory deficits and RBD. Moreover, cognitive symptoms were consistently reported at a lower prevalence in *LRRK2 G2019S* carriers with PD compared to non-carriers with idiopathic PD, which aligns with cross sectional studies using cognitive function tests.^40,41^ We showed that despite a longer disease duration, *LRRK2 G2019S* PD was associated with a similar prevalence of motor symptoms, including postural instability, suggesting a slower rate of progression. Taken together, our findings support the concept that the *LRRK2 G2019S* variant produces a milder form of parkinsonism with less impact to regions outside the substantia nigra (see Fig. 3), and aligns with observations in clinical cohorts.^13,42^ The differences in the clinical expression of PD in *LRRK2 G2019S* carriers compared to non-carriers with idiopathic PD may reflect differences in the core biology of *LRRK2 G2019S* PD, with α-synuclein aggregation in the CSF and Lewy body pathology at autopsy being absent in one third of *LRRK2* carriers with PD.^21^ This may be especially relevant as Lewy body pathology has been shown to have independent negative effects on global cognition and memory in longitudinal studies.^43^

Our findings have important implications for prodromal clinical trial designs in *LRRK2 G2019S* carriers. First, our results show that a high PRS confers an additional risk of PD in *LRRK2 G2019S* carriers, and may be useful as an enrichment stratification biomarker for candidate optimization.^44^ Second, we show that although not always absent, RBD and olfactory deficits are less prevalent in *LRRK2 G2019S* PD. This finding suggests that the current prodromal criteria that rely heavily on these non-motor features to predict the future likelihood of PD may underestimate the risk in *LRRK2 G2019S* carriers. Third, our phenoconversion rates to PD in *LRRK2 G2019S* carriers, although 10-fold higher than non-carriers, indicate large sample sizes would still be needed to adequately power clinical trials.^11^ This suggests that phenoconversion is unlikely to be a good primary outcome measure until we find a way to enrich the candidate section. Finally, we did not detect an increased prevalence of self-reported motor symptoms in older *LRRK2 G2019S* non-manifest carriers compared to matched non-carrier controls. Together, these findings underscore the need for wearable data endpoints that are sensitive to pick up sub-threshold parkinsonian symptoms in the early phases of neurodegeneration in high risk cohorts.

The geographic distribution of present-day *LRRK2 G2019S* carriers both aligns with the known history surrounding the dispersion of the variant while also revealing novel insights regarding the population structure.^3,45^ High carrier rates in North Africa support the concept of an ancient Moroccan Berber founder. The enriched geographic location in Eastern Europe—specifically in the area corresponding to the Pale of Settlement—aligns with the hypothesis of a subsequent flow of the *LRRK2 G2019S* variant into the Ashkenazi Jewish community, likely predating the 14^th^ century bottleneck.^3,4,45–47^

The discovery of carrier populations in the Latin Caribbean is novel. The Spanish colonization of these islands began in 1492, with the voyages of Columbus, which would introduce Iberian genetic ancestry into native populations. While Iberian ancestry is common among Puerto Rican and Cuban *LRRK2 G2019S* carriers in our cohort, North African and Ashkenazi ancestry was also present at lower frequencies in nearly all individuals. Historical records, previous ancestry analyses, and our findings align with the theory that the *LRRK2 G2019S* allele may have been introduced to the Caribbean by the male Sephardic Jewish Conversos who originally entered Spain via North Africa,^48^ and subsequently fled the Inquisitions, sailing to the new world as Conquistadors to become part of the founding populations.^49,50^ Mexican *LRRK2 G2019S* carriers, though sharing similar patterns of genetic ancestry with Latin Caribbean carriers, tended to have higher Ashkenazi ancestry with little North African ancestry. Jewish Conversos may have contributed to the introduction of the *LRRK2 G2019S* variant into Mexico, but the lack of North African ancestry suggests that the mutation may have originated primarily from the immigration of Yiddish-speaking Ashkenazim from eastern Europe following Mexico’s nineteenth-century Liberal reform, which included toleration of non-Catholic religions.^51^ Similarly, the influx of Eastern European Ashkenazi Jews who emigrated from the Pale of Settlement to South America likely also contributed to the high *LRRK2 G2019S* carrier rates that were observed in Uruguay, Argentina, and Brazil.^52^ Across Latin America, these founding populations remained geographically isolated, went through additional bottlenecks, followed by periods of population expansion, leading to a higher burden of rare variants.^53^ Ultimately, colonization, migration due to religious persecution, and other diasporas introduced the *LRRK2 G2019S* allele into populations outside of Europe and North Africa.

A major advantage of our study is the sample size of 23andMe’s *LRRK2 G2019S* cohort enrolled in this prospective study, which exceeds other published cohorts by nearly three-fold. Our findings in *LRRK2 G2019S* carriers are supported by a control group of over 2,000 non-carrier idiopathic PD patients and 100,000 controls, enabling comparisons with well powered age-matched groups. The phenotypic data collection points used in the study were collected through online self-reported answers that provide a unique insight into the patient’s “voice” and functional implications. These self-report measures indicated that motor symptoms, particularly tremor and postural instability, are the most prominent features of *LRRK2 G2019S* carriers with PD.

There are several limitations to consider. First, the cohort of *LRRK2 G2019S* carriers at 23andMe may be unique in some respects, as they are considered more affluent, educated, and perhaps more concerned about their health to seek out direct-to-consumer genetic testing. Nevertheless, they are assembled by virtue of them choosing direct-to-consumer genetic testing and thus not biased by clinical presentation within a neurological subspecialty as occurs in many prodromal cohorts. We relied on self-reported PD diagnosis, but have shown 100% concordance (*k*=1.00) with neurologist confirmed diagnoses in a validation study of 23andMe’s Parkinson’s Disease Community.^54^ Moreover, we reviewed information available in other surveys, which confirmed that the participants with PD reported being diagnosed by a neurologist (95.2%) or other physician (4.1%). 23andMe has demonstrated high validity with self-reported symptom measures.^55,56^ We are aware that RBD may be underreported in patients without a bed partner and some patients may be unaware that they have olfactory deficits, but this is unlikely to invalidate our findings as the prevalence of under-reporting is expected to be similar in carriers and non-carriers. There does, however, appear to be good concordance between self-reported motor symptoms of PD and findings on neurological examination.^57^ Compared to non-carriers with idiopathic PD, *LRRK2 G2019S* carriers with PD reported a 2-year earlier age of PD onset, and incident *LRRK2 G2019S* cases were diagnosed on average 6.6 years younger. It is possible that being aware of their *LRRK2 G2019S* carrier status prompted them to seek neurological care sooner, but it is also likely that the size of our cohort does reveal an earlier age of PD onset in *LRRK2 G2019S* that was not random. We saw the highest rates of follow-up in older *G2019S* carriers, which is why we focused our comparisons on older carriers >40 years of age who had the highest follow-up rates (see Fig. 1) and used education as a covariate in our survival analyses. Larger sample sizes are needed to determine whether the *LRRK2 G2019S* mutation offers some protection in dual carriers with a GBA N370S mutation, who do not appear to have an earlier onset of disease.^58^ Further work is needed to determine if the findings are generalizable to carriers of other *LRRK2* variants beyond *G2019S*.

The question remains as to why one-third of *LRRK2* PD patients do not show α-synuclein pathology at autopsy^19^ and have a negative α-synuclein seeding amplification assay test.^12^ This is important to understand as anti-α-synuclein based treatment strategies could potentially lack efficacy in a subset of *LRRK2 G2019S* carriers and/or these patients may not meet eligibility criteria to enter trials and test new therapies. However, this raises the bigger unanswered question of where *LRRK2 G2019S* carriers who meet clinical diagnostic criteria for PD, but lack evidence of α-synuclein seeding, fit in the new biological definitions of PD that are likely to emerge, especially as causal role of α-synuclein aggregates in PD is still being debated.^59,60^

In summary, this study shows that *LRRK2 G2019S* PD is associated with fewer self-reported cognitive and olfactory deficits and a lower prevalence of RBD. The findings may have implications for the design of early interventional trials in *LRRK2 G2019S* carriers, including the need for more precise prodromal clinical criteria to detect early disease onset, using high PRS scores as an enrichment biomarker to optimize candidate selection for a high yield of phenoconverters, and developing more sensitive endpoints for tracking this slow progressing mild-motor subtype of PD. Finally, our ancestry results showing high carrier rates throughout the Latin Caribbean and South America may be helpful for selecting populations in other countries for genetic screening as ways to advance clinical trial recruitment in *LRRK2 G2019S* carriers.

## Data Availability

Model outputs for all logistic regressions are provided as Supplementary Materials. Individual-level data are not publicly available due to participant confidentiality, and in accordance with the IRB-approved protocol under which the study was conducted. No custom code or software was generated as part of the study. Details of all software packages used for data processing and/or analysis may be found in the Methods.

## Acknowledgements

We would like to thank Mina Kmiecik for data illustrations, and the research participants and employees of 23andMe for making this work possible.

## Funding

We thank The Michael J. Fox Foundation for Parkinson’s Research for funding this research (MJK, KS, and LNK).

## Competing Interests

At the time of their contributions, authors were employed by and held stock or stock options in 23andMe, Inc.

## Appendix

The following members of the 23andMe Research Team contributed to this study:

Adam Auton, Elizabeth Babalola, Robert K. Bell, Jessica Bielenberg, Johnathan Bowes, Katarzyna Bryc, Ninad S. Chaudhary, Sayantan Das, Emily DelloRusso, Sarah L. Elson, Nicholas Eriksson, Will Freyman, Julie M. Granka, Alejandro Hernandez, Barry Hicks, Ethan M. Jewett, Yunxuan Jiang, Katelyn Kukar, Alan Kwong, Keng-Han Lin, Bianca A. Llamas, Maya Lowe, Matthew H. McIntyre, Meghan E. Moreno, Priyanka Nandakumar, Dominique T. Nguyen, Jared O’Connell, Aaron A. Petrakovitz, G. David Poznik, Alexandra Reynoso, Morgan Schumacher, Leah Selcer, Anjali J. Shastri, Qiaojuan Jane Su, Susana A. Tat, Vinh Tran, Xin Wang, Wei Wang, Catherine H. Weldon, Peter Wilton, Corinna D. Wong

## Notes

### Author Declarations

All study research participants were >18 years old, US residents, and provided informed consent to volunteer to participate (protocol approval: AAHRPP-accredited Salus IRB).

## References

1. Simpson C, Vinikoor-Imler L, Nassan FL, et al. Prevalence of ten LRRK2 variants in Parkinson’s disease: A comprehensive review. Parkinsonism Relat Disord. 2022;98:103–113. doi:10.1016/j.parkreldis.2022.05.012

2. Biskup S, West AB. Zeroing in on LRRK2-linked pathogenic mechanisms in Parkinson’s disease. Biochim Biophys Acta BBA - Mol Basis Dis. 2009;1792(7):625–633. doi:10.1016/j.bbadis.2008.09.015

3. Haj RBE, Salmi A, Regragui W, et al. Evidence for prehistoric origins of the G2019S mutation in the North African Berber population. PLOS ONE. 2017;12(7):e0181335. doi:10.1371/journal.pone.0181335

4. Bar-Shira A, Hutter CM, Giladi N, Zabetian CP, Orr-Urtreger A. Ashkenazi Parkinson’s disease patients with the LRRK2 G2019S mutation share a common founder dating from the second to fifth centuries. neurogenetics. 2009;10(4):355-358. doi:10.1007/s10048-009-0186-0

5. Alessi DR, Sammler E. LRRK2 kinase in Parkinson’s disease. Science. 2018;360(6384):36-37. doi:10.1126/science.aar5683

6. Gasper R, Meyer S, Gotthardt K, Sirajuddin M, Wittinghofer A. It takes two to tango: regulation of G proteins by dimerization. Nat Rev Mol Cell Biol. 2009;10(6):423–429. doi:10.1038/nrm2689

7. Nguyen APT, Tsika E, Kelly K, et al. Dopaminergic neurodegeneration induced by Parkinson’s disease-linked G2019S LRRK2 is dependent on kinase and GTPase activity. Proc Natl Acad Sci. 2020;117(29):17296–17307. doi:10.1073/pnas.1922184117

8. Di Maio R, Hoffman EK, Rocha EM, et al. LRRK2 activation in idiopathic Parkinson’s disease. Sci Transl Med. 2018;10(451):eaar5429. doi:10.1126/scitranslmed.aar5429

9. Trinh J, Amouri R, Duda JE, et al. A comparative study of Parkinson’s disease and leucine-rich repeat kinase 2 p.G2019S parkinsonism. Neurobiol Aging. 2014;35(5):1125-1131. doi:10.1016/j.neurobiolaging.2013.11.015

10. Jensen-Roberts S, Myers TL, Auinger P, et al. A Remote Longitudinal Observational Study of Individuals at Genetic Risk for Parkinson Disease: Baseline Results. Neurol Genet. 2022;8(5). doi:10.1212/NXG.0000000000200008

11. Joza S, Hu MT, Jung KY, et al. Progression of clinical markers in prodromal Parkinson’s disease and dementia with Lewy bodies: a multicentre study. Brain. 2023;146(8):3258–3272. doi:10.1093/brain/awad072

12. Siderowf A, Concha-Marambio L, Lafontant DE, et al. Assessment of heterogeneity among participants in the Parkinson’s Progression Markers Initiative cohort using α-synuclein seed amplification: a cross-sectional study. Lancet Neurol. 2023;22(5):407–417. doi:10.1016/S1474-4422(23)00109-6

13. Simuni T, Merchant K, Brumm MC, et al. Longitudinal clinical and biomarker characteristics of non-manifesting LRRK2 G2019S carriers in the PPMI cohort. Npj Park Dis. 2022;8(1):140. doi:10.1038/s41531-022-00404-w

14. Bestwick JP, Auger SD, Simonet C, et al. Improving estimation of Parkinson’s disease risk—the enhanced PREDICT-PD algorithm. Npj Park Dis. 2021;7(1):1–7. doi:10.1038/s41531-021-00176-9

15. Heinzel S, Berg D, Gasser T, et al. Update of the MDS research criteria for prodromal Parkinson’s disease. Mov Disord. 2019;34(10):1464–1470. doi:10.1002/mds.27802

16. Pont-Sunyer C, Iranzo A, Gaig C, et al. Sleep Disorders in Parkinsonian and Nonparkinsonian LRRK2 Mutation Carriers. PLOS ONE. 2015;10(7):e0132368. doi:10.1371/journal.pone.0132368

17. Vilas D, Tolosa E, Quintana M, et al. Olfaction in LRRK2 Linked Parkinson’s Disease: Is It Different from Idiopathic Parkinson’s Disease? J Park Dis. 2020;10(3):951–958. doi:10.3233/JPD-201972

18. Saunders-Pullman R, Mirelman A, Alcalay RN, et al. Progression in the LRRK2-Associated Parkinson Disease Population. JAMA Neurol. 2018;75(3):312–319. doi:10.1001/jamaneurol.2017.4019

19. Kalia LV, Lang AE, Hazrati LN, et al. Clinical Correlations With Lewy Body Pathology in LRRK2-Related Parkinson Disease. JAMA Neurol. 2015;72(1):100–105. doi:10.1001/jamaneurol.2014.2704

20. Lanore A, Casse F, Tesson C, et al. Differences in Survival across Monogenic Forms of Parkinson’s Disease. Ann Neurol. 2023;94(1):123–132. doi:10.1002/ana.26636

21. Attems J, Toledo JB, Walker L, et al. Neuropathological consensus criteria for the evaluation of Lewy pathology in post-mortem brains: a multi-centre study. Acta Neuropathol (Berl). 2021;141(2):159–172. doi:10.1007/s00401-020-02255-2

22. 23andMe, Inc. 23andMe® Personal Genome Service® (PGS) Package Insert. Published online 2023. doi:https://permalinks.23andme.com/pdf/package_insert_v5.pdf?_gl=1*11q2npm*_ga*MjM4NTAxOTMxLjE2OTI4MTc3MTM.*_ga_G330GF3ZFF*MTY5MjgxNzcxMi4xLjEuMTY5MjgxNzgyMi4wLjAuMA.

23. Kroenke K, Spitzer RL, Williams JBW. The PHQ-9. J Gen Intern Med. 2001;16(9):606–613. doi:10.1046/j.1525-1497.2001.016009606.x

24. Bondy SJ, Victor JC, Diemert LM. Origin and use of the 100 cigarette criterion in tobacco surveys. Tob Control. 2009;18(4):317–323. doi:10.1136/tc.2008.027276

25. Ho D, Imai K, King G, Stuart EA. MatchIt: Nonparametric Preprocessing for Parametric Causal Inference. J Stat Softw. 2011;42:1–28. doi:10.18637/jss.v042.i08

26. Benjamini Y, Hochberg Y. Controlling the False Discovery Rate: A Practical and Powerful Approach to Multiple Testing. J R Stat Soc Ser B Methodol. 1995;57(1):289–300. doi:10.1111/j.2517-6161.1995.tb02031.x

27. Freyman WA, McManus KF, Shringarpure SS, et al. Fast and Robust Identity-by-Descent Inference with the Templated Positional Burrows–Wheeler Transform. Mol Biol Evol. 2021;38(5):2131–2151. doi:10.1093/molbev/msaa328

28. Traag VA, Waltman L, van Eck NJ. From Louvain to Leiden: guaranteeing well-connected communities. Sci Rep. 2019;9(1):5233. doi:10.1038/s41598-019-41695-z

29. Durand EY, Do CB, Mountain JL, Macpherson JM. Ancestry Composition: A Novel, Efficient Pipeline for Ancestry Deconvolution. Published online October 18, 2014:010512. doi:10.1101/010512

30. Iacus SM, King G, Porro G. cem: Software for Coarsened Exact Matching. J Stat Softw. 2009;30(9). doi:10.18637/jss.v030.i09

31. Braak H, Ghebremedhin E, Rüb U, Bratzke H, Del Tredici K. Stages in the development of Parkinson’s disease-related pathology. Cell Tissue Res. 2004;318(1):121–134. doi:10.1007/s00441-004-0956-9

32. Chiaro G, Calandra-Buonaura G, Cecere A, et al. REM sleep behavior disorder, autonomic dysfunction and synuclein-related neurodegeneration: where do we stand? Clin Auton Res. 2018;28(6):519–533. doi:10.1007/s10286-017-0460-4

33. Nalls MA, Blauwendraat C, Vallerga CL, et al. Identification of novel risk loci, causal insights, and heritable risk for Parkinson’s disease: a meta-analysis of genome-wide association studies. Lancet Neurol. 2019;18(12):1091–1102. doi:10.1016/S1474-4422(19)30320-5

34. Chang D, Nalls MA, Hallgrímsdóttir IB, et al. A meta-analysis of genome-wide association studies identifies 17 new Parkinson’s disease risk loci. Nat Genet. 2017;49(10):1511–1516. doi:10.1038/ng.3955

35. Fahed AC, Wang M, Homburger JR, et al. Polygenic background modifies penetrance of monogenic variants for tier 1 genomic conditions. Nat Commun. 2020;11(1):3635. doi:10.1038/s41467-020-17374-3

36. R Core Team. R: A language and environment for statistical computing. Published online 2020. https://www.R-project.org/

37. Therneau T. A Package for Survival Analysis in R. Published online 2020. https://CRAN.R-project.org/package=survival

38. Jackson C. flexsurv: A Platform for Parametric Survival Modeling in R. J Stat Softw. 2016;70:1-33. doi:10.18637/jss.v070.i08

39. Wickham H. Ggplot2: Elegant Graphics for Data Analysis. Springer-Verlag; 2016.

40. Ben Romdhan S, Farhat N, Nasri A, et al. LRRK2 G2019S Parkinson’s disease with more benign phenotype than idiopathic. Acta Neurol Scand. 2018;138(5):425–431. doi:10.1111/ane.12996

41. Srivatsal S, Cholerton B, Leverenz JB, et al. Cognitive profile of LRRK2-related Parkinson’s disease. Mov Disord. 2015;30(5):728–733. doi:10.1002/mds.26161

42. Myers TL, Augustine EF, Baloga E, et al. Recruitment for Remote Decentralized Studies in Parkinson’s Disease. J Park Dis. 2022;12(1):371–380. doi:10.3233/JPD-212935

43. Palmqvist S, Rossi M, Hall S, et al. Cognitive effects of Lewy body pathology in clinically unimpaired individuals. Nat Med. 2023;29(8):1971–1978. doi:10.1038/s41591-023-02450-0

44. Liu G, Peng J, Liao Z, et al. Genome-wide survival study identifies a novel synaptic locus and polygenic score for cognitive progression in Parkinson’s disease. Nat Genet. 2021;53(6):787–793. doi:10.1038/s41588-021-00847-6

45. Waldman S, Backenroth D, Harney É, et al. Genome-wide data from medieval German Jews show that the Ashkenazi founder event pre-dated the 14th century. Cell. 2022;185(25):4703–4716.e16. doi:10.1016/j.cell.2022.11.002

46. Campbell CL, Palamara PF, Dubrovsky M, et al. North African Jewish and non-Jewish populations form distinctive, orthogonal clusters. Proc Natl Acad Sci. 2012;109(34):13865–13870. doi:10.1073/pnas.1204840109

47. Ostrer H, Skorecki K. The population genetics of the Jewish people. Hum Genet. 2013;132(2):119–127. doi:10.1007/s00439-012-1235-6

48. Adams SM, Bosch E, Balaresque PL, et al. The Genetic Legacy of Religious Diversity and Intolerance: Paternal Lineages of Christians, Jews, and Muslims in the Iberian Peninsula. Am J Hum Genet. 2008;83(6):725–736. doi:10.1016/j.ajhg.2008.11.007

49. Fortes-Lima C, Bybjerg-Grauholm J, Marin-Padrón LC, et al. Exploring Cuba’s population structure and demographic history using genome-wide data. Sci Rep. 2018;8(1):11422. doi:10.1038/s41598-018-29851-3

50. Mooney JA, Huber CD, Service S, et al. Understanding the Hidden Complexity of Latin American Population Isolates. Am J Hum Genet. 2018;103(5):707–726. doi:10.1016/j.ajhg.2018.09.013

51. Krause CA. Mexico—Another Promised Land? A Review of Projects for Jewish Colonization in Mexico: 1881–1925. Am Jew Hist Q. 1972;61(4):325-341.

52. Norcliffe-Kaufmann L, Slaugenhaupt SA, Kaufmann H. Familial dysautonomia: History, genotype, phenotype and translational research. Prog Neurobiol. 2017;152:131–148. doi:10.1016/j.pneurobio.2016.06.003

53. Browning SR, Browning BL, Daviglus ML, et al. Ancestry-specific recent effective population size in the Americas. PLOS Genet. 2018;14(5):e1007385. doi:10.1371/journal.pgen.1007385

54. Dorsey ER, Darwin KC, Mohammed S, et al. Virtual research visits and direct-to-consumer genetic testing in Parkinson’s disease. Digit Health. 2015;1:2055207615592998. doi:10.1177/2055207615592998

55. Francke U, Naughton B, Mountain J, et al. Efficient Replication of Over 180 Genetic Associations with Self-Reported Medical Data. Nat Preced. Published online June 7, 2011:1–1. doi:10.1038/npre.2011.6014.2

56. Winslow AR, Hyde CL, Wilk JB, et al. Self-report data as a tool for subtype identification in genetically-defined Parkinson’s Disease. Sci Rep. 2018;8(1):12992. doi:10.1038/s41598-018-30843-6

57. Lieberman A, Krishnamurthi N, Dhall R, et al. A Simple Question About Falls to Distinguish Balance and Gait Difficulties in Parkinson’s Disease. Int J Neurosci. 2012;122(12):710–715. doi:10.3109/00207454.2012.711399

58. Ortega RA, Wang C, Raymond D, et al. Association of Dual LRRK2 G2019S and GBA Variations With Parkinson Disease Progression. JAMA Netw Open. 2021;4(4):e215845. doi:10.1001/jamanetworkopen.2021.5845

59. Chopra A, Outeiro TF. Aggregation and beyond: alpha-synuclein-based biomarkers in synucleinopathies. Brain. Published online August 1, 2023:awad260. doi:10.1093/brain/awad260

60. Ezzat K, Sturchio A, Espay AJ. Chapter 3 - The shift to a proteinopenia paradigm in neurodegeneration. In: Espay AJ, ed. Handbook of Clinical Neurology. Vol 193. Precision Medicine in Neurodegenerative Disorders, Part II. Elsevier; 2023:23-32. doi:10.1016/B978-0-323-85555-6.00001-1

