## Supplementary Materials for "Genetic Analysis and Natural History of Parkinson’s Disease Due to the LRRK2 G2019S Variant"

### Supplementary Material

#### Genotyping

The V1 and V2 platforms were variants of the Illumina HumanHap550+ BeadChip and contained a total of approximately 560,000 SNPs, including about 25,000 custom SNPs selected by 23andMe. The V3 platform was based on the Illumina OmniExpress + BeadChip and contained a total of about 950,000 SNPs and custom content to improve the overlap with our V2 array. The V4 platform is a fully custom array and includes a lower redundancy subset of V2 and V3 SNPs with additional coverage of lower-frequency coding variation, and about 570,000 SNPs. The V5 platform is an Illumina Infinium Global Screening Array of about 640,000 SNPs supplemented with about 50,000 SNPs of custom content.

#### Imputation Reference Panels

Variants were imputed using a reference panel comprised of three other reference panels: the publicly available Human Reference Consortium (HRC) and UK BioBank (UKBB) 200K Whole Exome Sequencing (WES) reference panels, and the 23andMe reference panel. HRC data were downloaded from the European Genome-Phenome Archive at the European Bioinformatics Institute (accession EGAD00001002729). Variants were converted to hg38 and excluded if their new positions were on a different chromosome. Variants were then re-phased using SHAPEIT4 (https://odelaneau.github.io/shapeit4/). Finally, singletons were excluded. The final HRC reference panel included 27,165 samples and 39,057,040 SNPs (no indels).

The UKBB 200K WES reference panel was built following a recently published approach (Barton et al., 2021). UKBB genotyping data included 486,633 samples and 681,298 variants, and was lifted from hg19 to hg38 and re-phased using SHAPEIT4. UKBB 200K WES data included 200,643 whole exome sequenced samples (mostly British ancestry) and more than 22M variants (https://www.ukbiobank.ac.uk/media/cfulxh52/uk-biobank-exome-release-faq_v9-december-2020.pdf). Multi-allelic variants were split into bi-allelic variants using bcftools. Genotypes with GQ<20 were set to missing. Variants with >20% missingness, a minor allele count of 0, or an inbreeding coefficient < -0.3 (high heterozygosity) were removed. After QC, 17,975,023 UKBB WES variants remained. For the 199,815 samples with both WES and genotype array data, the variants were merged and re-phased using SHAPEIT4. After excluding singletons, the UKBB 200K WES reference panel included 199,815 samples and 17,082,588 variants (16,115,376 SNPs and 967,212 indels).

The 23andMe reference panel included 12,217 samples from: consented 23andMe customers, the 1000 Genomes Project (Auton et al., 2015), Syndip (Li et al., 2018), the Genotype-Tissue Expression (GTEx) project v8 (Aguet et al., 2019), the Human Genome Diversity Project (HGDP) (Bergström et al., 2020), and the Simons Genome Diversity Project (SGDP) (Mallick et al., 2016). All samples (except for those from GTEx v8) were consented for research as of 2020-01-18, had sufficient depth of coverage (greater than the median - 3*MAD (median absolute deviation) within each cohort), had contamination < 0.05 as estimated by *verifybamid*, had <0.05 chimeric reads, had median insert size >= 250 bp, had r2 with genotyping array >= 0.8, and had >= 3M SNPs called. GTEx v8 samples were sometimes slightly outside these bounds, but were included due to their value in eQTL mapping. All samples were aligned to GRCh38_full_analysis_set_plus_decoy_hla.fa9 and duplicate marked. Datasets sequenced before 01/01/2019 were re-aligned using an in-house pipeline consisting of bwa mem 0.7.15-r1140 alignment and duplicate marking with samblaster v0.1.24. Datasets sequenced after 01/01/2019 were processed using a well-known public pipeline from the Broad Institute (Aguet et al., 2019) that combined bwa mem 0.7.15-r1140 alignment, Picard MarkDuplicates 2.15.0, and BQSR with GATK 4.beta.5. Variants were called in each individual sample using DeepVariant-0.8.0 11 to produce GVCFs. The GVCFs were then joint-called using GLnexus-1.2.3 12. Singletons were removed, genotypes with GQ<20 were set to missing, variants with >20% missingness (after the GQ filter) were removed, and variants with >30% excess heterozygosity were removed. Finally, variants were phased using SHAPEIT4. SHAPEIT4 also imputed missing genotypes and produced a final panel without missingness. The final 23andMe reference panel included 12,217 samples and 82,078,539 variants (73,852,355 SNPs + 8,226,184 indels).

#### Imputation

Imputation was performed using Beagle 5 (Browning et al., 2018). Because participants were genotyped on one of the five genotyping platforms (v1 to v5), imputation was performed independently for each platform. Similarly, participants were classified into one of five ancestry-based populations as described previously (https://www.23andme.com/ancestry-composition-guide/) and imputation was performed independently for each population. We used a two-step imputation process: first combining results from the HRC and 23andMe reference panels, then combining the HRC+23andMe results with UKBB 200K WES results. In the first step, there were 85,099,656 variants found in either the HRC or 23andMe reference panels. HRC-specific variants (n=3,021,117) were imputed using all HRC reference panel samples. 23andMe-specific variants (n=46,042,616) were imputed using all 23andMe reference panel samples. Variants found in both HRC and 23andMe reference panels (hereafter referred to as HRC+23andMe, n= 36,035,923) were imputed using all HRC+23andMe reference panel samples. In the second step, there were 99,675,338 variants found in either the HRC+23andMe or UKBB 200K WES reference panels. HRC+23andMe-specific variants were imputed as described in step 1 (n=82,592,750). UKBB 200K WES-specific variants were imputed using the UKBB 200K WES reference panel (n= 14,575,682). Variants found in both the HRC+23andMe and the UKBB 200K WES reference panels (n= 2,506,906) were imputed in each panel separately and the result with the higher imputation quality was used. Imputation quality was taken as the sample size-weighted imputation R2 value across genotyping platforms and populations.

#### Polygenic Risk Score Calculation

Both the original and modified PRS were a weighted sum of risk allele counts for the included SNPs that were matched using CPRA (chromosome, position, reference allele, alternative allele) format and harmonized to 23andMe imputation panel.[35] We removed SNPs with imputation $R^2$<0.5 or a difference in MAF>30% between the Nalls et al.[33] and 23andMe variants. These quality control procedures removed an additional 69 SNPs from both PRS (67 unmatched and 2 low $R^2$ variants) resulting in a total of 1,770 and 1,724 variants for the original and modified PRS, respectively.

#### Principal Components of Ancestry

PCs were derived via principal component analysis performed on one million randomly selected participants from 23andMe's Research Cohort using 63,528 high quality genotyped variants present across all five genotyping platforms. Loadings for participants not included in the analysis were obtained by projection, combining the eigenvectors of the analysis and the SNP weights.

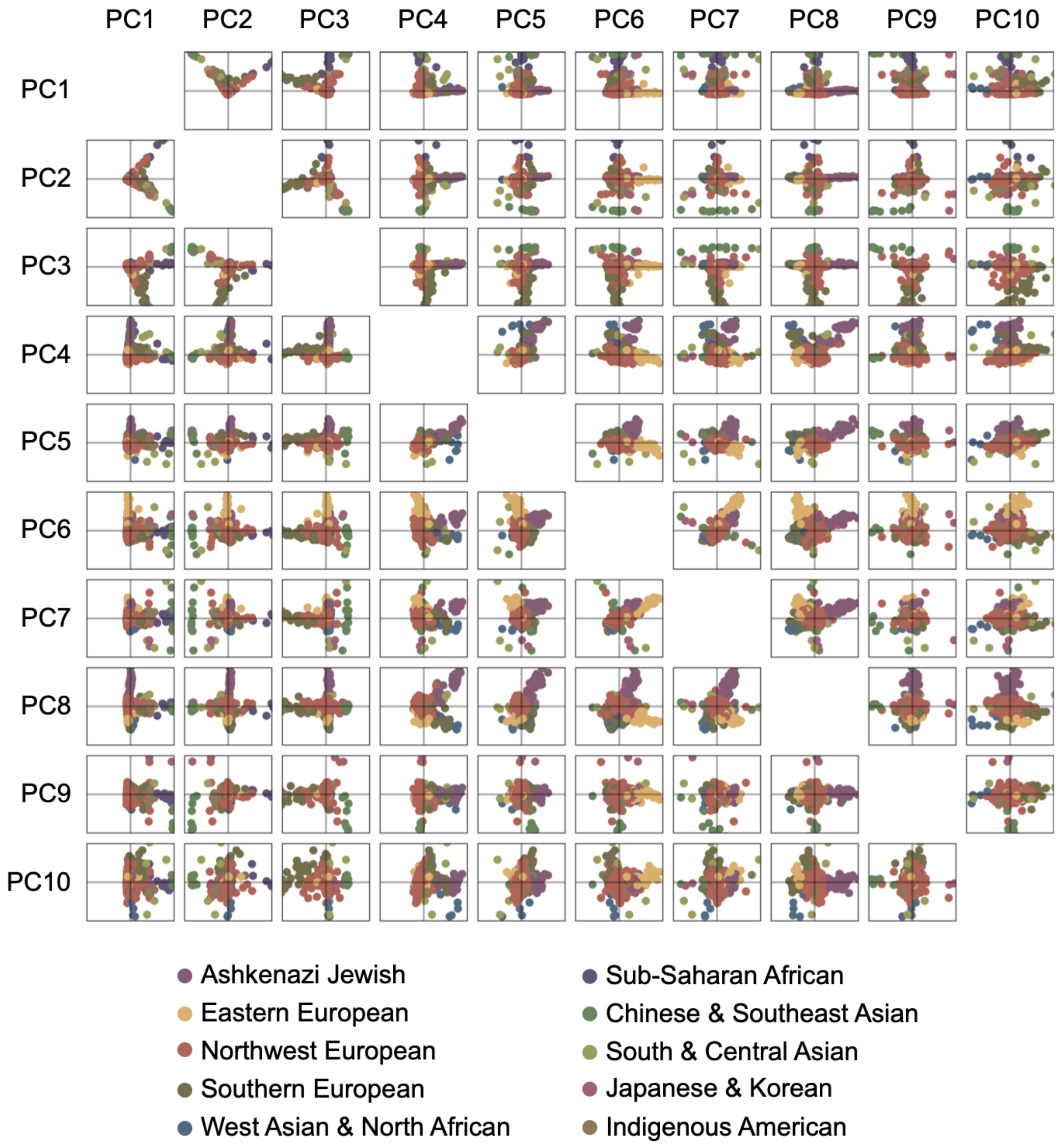


**Supplementary Figure 1.** Scatter matrix of the first 10 ancestry principal components (PCs) plotted from a random sampling of *n*=1,000 PIP participants. We classified individuals by their maximum genetic ancestry using 12 regional populations predicted by Ancestry Composition (Durand et al., 2014). 0.03% of participants with ancestry that is rare in the PIP cohort (i.e., Oceanian or Northern Asian) were excluded in this random sample and are therefore not visualized.

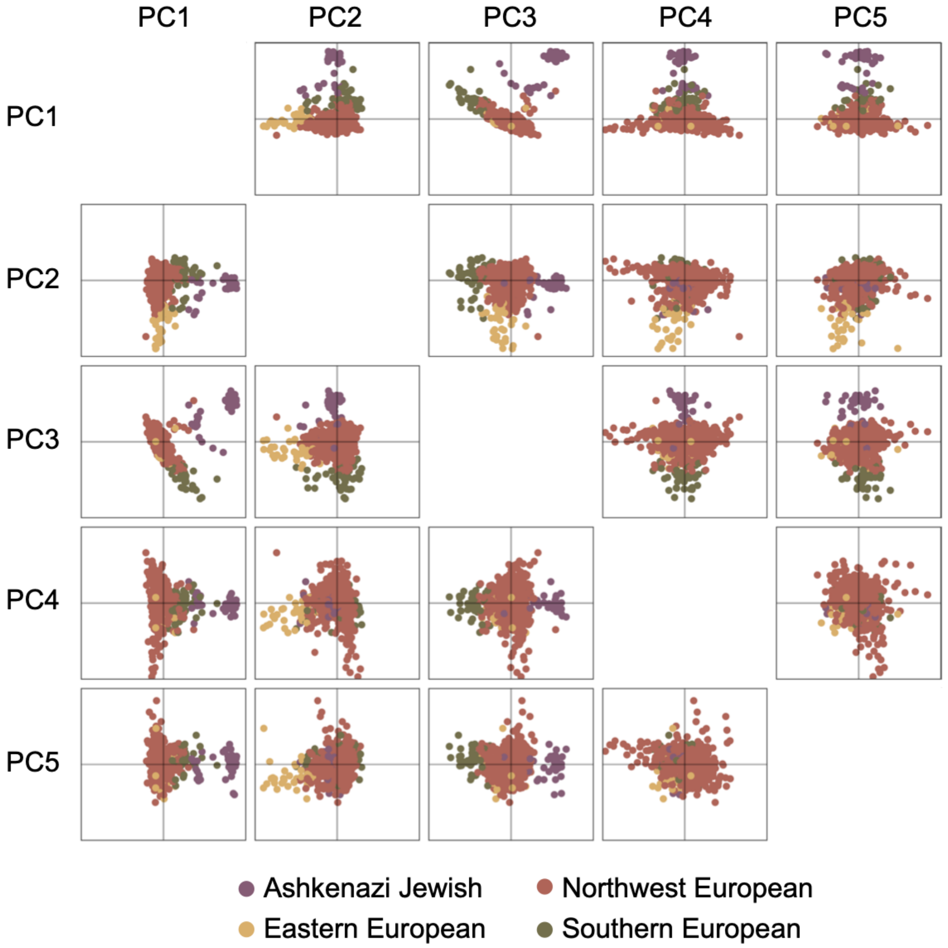


**Supplementary Figure 2.** Scatter matrix of the first 5 ancestry principal components (PCs) for European participants plotted from a random sampling of *n*=1,000 PIP participants. We classified individuals by their maximum genetic ancestry using 12 regional populations predicted by Ancestry Composition (Durand et al., 2014); however, only European participants are plotted here.

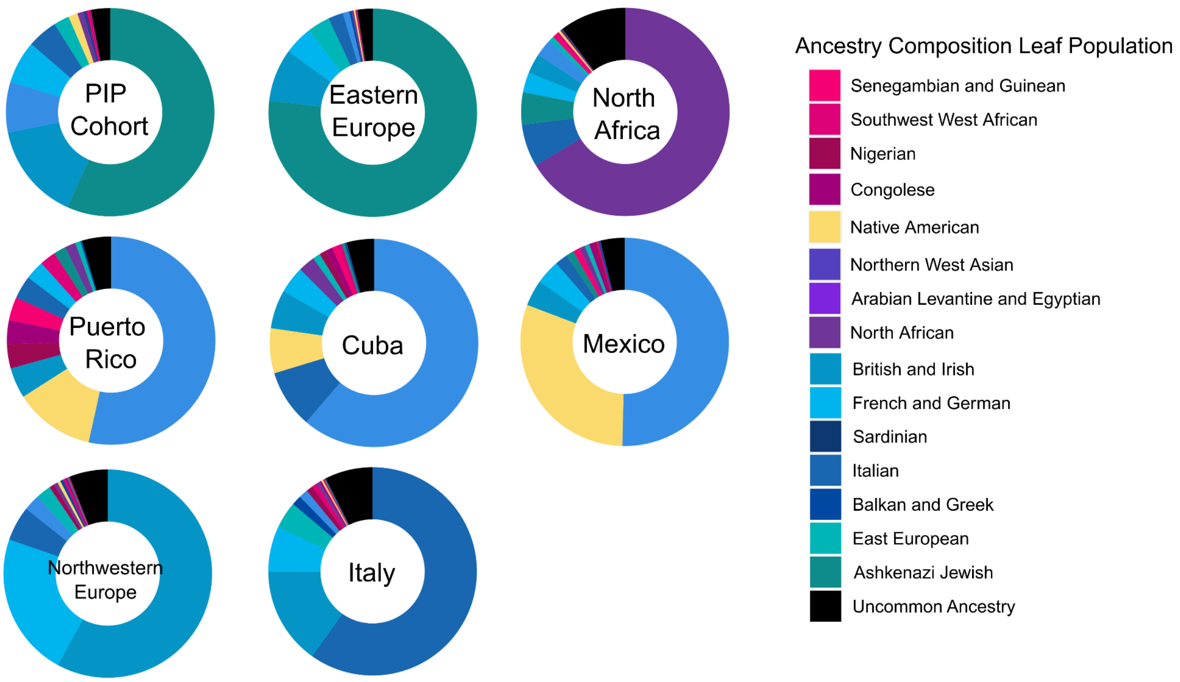


**Supplementary Figure 3. Mean Ancestry Composition of the PIP cohort and LRRK2 G2019S carrier genetic groups.** Averages of 23andMe Ancestry Composition populations that are present at a rate of $\geq$ 5% in the LRRK2 G2019S cohort shown in the entire PIP Cohort and each of the genetic groups determined from the entire LRRK2 G2019s cohort. All uncommon ancestry populations are binned into the broader "Uncommon Ancestry" category.

Supplementary Table 1. *Clinical Characteristics of the Parkinson's Impact Project Cohort at Study Entry: Additional Groups.*

| **Category** | **Measure** | **All LRRK2 G2019S** | **All Non-carriers** | **Non-carrier Controls Matched to PD Participants** |
|---|---|---|---|---|
| Demographics | N | 1286 | 109472 | 23010 |
| | Male | 40% | 37% | 53% |
| | Age (years) | 54.32 (0.45) | 54.81 (0.05) | 69 (0.06) |
| | BMI | 27.04 (0.17) | 28.31 (0.02) | 28.32 (0.04) |
| | European | 80% | 83% | 89% |
| | Ashkenazi | 43% | 2% | 4% |
| | GBA N370S | 3% | 1% | 1% |
| | Education | 83% | 66% | 69% |
| Motor | Tremor | 21% | 11% | 11% |
| | Imbalance | 26% | 21% | 25% |
| | Bradykinesia | 15% | 10% | 15% |
| | Reduced Arm Swing | 16% | 7% | 8% |
| Prodromal Markers | RBD | 5% | 3% | 2% |
| | MCI Diagnosis | 2% | 1% | 2% |
| | Depression (PHQ^) | 4.56 (0.13) | 4.3 (0.01) | 3.01 (0.03) |
| | Olfactory Difficulties | 9% | 8% | 9% |
| | OH | 33% | 27% | 22% |
| | Constipation | 5% | 5% | 4% |
| | Urinary Difficulties | 26% | 21% | 23% |
| | Erectile Dysfunction | 24% | 25% | 31% |
| | Daytime Somnolence | 13% | 13% | 8% |
| Risk Factors | Occupational Toxins | 2% | 5% | 6% |
| | Occupational PPE | 27% | 36% | 34% |
| | Non-smoking | 63% | 59% | 52% |
| | Non-use of Caffeine | 26% | 28% | 25% |
| | Physical Inactivity | 6% | 8% | 8% |
| | 1° Relative with PD | 27% | 6% | 8% |
| | Head Injury (with LOC) | 11% | 13% | 14% |

*Note.* Continuous measures are presented *M* (*SEM*); ^PHQ-9 was administered without question #9; PD=Parkinson's Disease; BMI=Body Mass Index; RBD=REM Sleep Behavior Disorder; MCI=Mild Cognitive Impairment; OH=Orthostatic Hypotension; PPE=Personal Protective Equipment; LOC=Loss of Consciousness; FDR=False Discovery Rate.

Supplementary Table 2. *Sample Sizes of Grandparent Birth Countries by Cohort.*

| Grandparent Birth Country | PIP cohort count | G2019S cohort count | Total participant count |
|---|---|---|---|
| United States of America (US) | 1338 | 6846 | 8650108 |
| Russian Federation (RU) | 322 | 1377 | 170338 |
| Poland (PL) | 297 | 1251 | 240349 |
| Ukraine (UA) | 172 | 855 | 124546 |
| Puerto Rico (PR) | 72 | 823 | 171435 |
| Mexico (MX) | 45 | 700 | 508441 |
| Italy (IT) | 118 | 697 | 424453 |
| Cuba (CU) | 79 | 533 | 94964 |
| Great Britain (GB) | 63 | 473 | 664248 |
| Belarus (BY) | 72 | 410 | 35353 |
| Lithuania (LT) | 93 | 379 | 47787 |
| Germany (DE) | 93 | 372 | 281918 |
| Morocco (MA) | 5 | 283 | 13235 |
| Spain (ES) | 39 | 238 | 98614 |
| Austria (AT) | 65 | 233 | 47368 |
| Hungary (HU) | 35 | 213 | 65335 |
| Romania (RO) | 40 | 212 | 55645 |
| Algeria (DZ) | 11 | 209 | 8224 |
| Portugal (PT) | 9 | 151 | 63549 |
| Canada (CA) | 38 | 142 | 215170 |
| Brazil (BR) | 27 | 142 | 69939 |
| Latvia (LV) | 34 | 125 | 14559 |
| France (FR) | 7 | 112 | 88628 |
| Colombia (CO) | 22 | 107 | 73827 |
| Ireland (IE) | 19 | 106 | 201917 |
| Israel (IL) | 11 | 88 | 10455 |
| Dominican Republic (DO) | 18 | 87 | 55910 |
| Chile (CL) | 12 | 84 | 18936 |
| Tunisia (TN) | 5 | 79 | 4705 |
| Venezuela (VE) | 9 | 72 | 25486 |
| Greece (GR) | 16 | 61 | 69298 |
| Argentina (AR) | 5 | 60 | 27097 |
| Egypt (EG) | 7 | 60 | 18312 |
| Guatemala (GT) | 5 | 56 | 30428 |
| Moldova (MD) | 5 | 54 | 7867 |
| Peru (PE) | 16 | 53 | 35662 |
| Czechia (CZ) | 9 | 53 | 37483 |
| Syrian Arab Republic (SY) | 5 | 49 | 16185 |
| Turkey (TR) | 5 | 49 | 45258 |
| South Africa (ZA) | 7 | 48 | 26125 |
| Norway (NO) | 6 | 42 | 58641 |
| Australia (AU) | 5 | 37 | 44753 |
| Sweden (SE) | 7 | 37 | 63928 |
| Slovakia (SK) | 5 | 32 | 26805 |
| Lebanon (LB) | 5 | 31 | 26321 |
| Switzerland (CH) | 8 | 28 | 29085 |
| Philippines (PH) | 5 | 25 | 106280 |
| Netherlands (NL) | 5 | 24 | 46634 |
| Honduras (HN) | 5 | 22 | 16703 |
| Iran (IR) | 5 | 18 | 46936 |

| | | | |
|---|---|---|---|
| Uruguay (UY) | 5 | 17 | 5835 |
| Ecuador (EC) | 5 | 15 | 29453 |
| Iraq (IQ) | 5 | 15 | 13244 |
| China (CN) | 6 | 14 | 129838 |
| Panama (PA) | 7 | 13 | 12510 |
| Palestine (PS) | 5 | 13 | 10489 |
| Jamaica (JM) | 5 | 11 | 42182 |
| Nicaragua (NI) | 5 | 11 | 19022 |
| Jordan (JO) | 5 | 11 | 3263 |
| Bulgaria (BG) | 5 | 11 | 17555 |
| India (IN) | 5 | 10 | 109533 |
| Zimbabwe (ZW) | 5 | 10 | 1786 |
| Iceland (IS) | 5 | 10 | 3830 |
| Uzbekistan (UZ) | 5 | 10 | 5907 |
| Cyprus (CY) | 5 | 10 | 9802 |
| North Macedonia (MK) | 5 | 10 | 6109 |
| Kenya (KE) | 5 | 10 | 2469 |
| Estonia (EE) | 5 | 10 | 5212 |

*Note.* Reported grandparent birth countries in the PIP cohort, the full LRRK2 G2019S carrier cohort, which includes the PIP cohort, and all research participants in the 23andMe database that reported their grandparents were born in the same geographic regions. To protect participant privacy, we excluded countries with small counts ($n$<5).

Supplementary Table 3. *Reported Grandparent Birth Countries by LRRK2 G2019S Genetic Groups.*

| Genetic group | Grandparent birth country | G2019S carrier count | Total participant count | Carrier frequency |
|---|---|---|---|---|
| Cuba | Cuba (CU) | 474 | 95369 | 0.00497 |
| Cuba | Chile (CL) | 71 | 19036 | 0.00373 |
| Cuba | Venezuela (VE) | 57 | 25653 | 0.00222 |
| Cuba | Portugal (PT) | 128 | 63740 | 0.00201 |
| Cuba | Uruguay (UY) | 10 | 5859 | 0.00171 |
| Cuba | Brazil (BR) | 111 | 70276 | 0.00158 |
| Cuba | Spain (ES) | 120 | 98925 | 0.00121 |
| Cuba | Dominican Republic (DO) | 65 | 56217 | 0.00116 |
| Cuba | Colombia (CO) | 79 | 74264 | 0.00106 |
| Cuba | Argentina (AR) | 17 | 27224 | 0.00062 |
| Cuba | Italy (IT) | 72 | 425699 | 0.00017 |
| Cuba | France (FR) | 13 | 88880 | 0.00015 |
| Cuba | Germany (DE) | 10 | 282663 | 0.00004 |
| Cuba | United States of America (US) | 189 | 8681625 | 0.00002 |
| Cuba | Great Britain (GB) | 11 | 666326 | 0.00002 |
| East Europe | Belarus (BY) | 407 | 35494 | 0.01147 |
| East Europe | Latvia (LV) | 123 | 14602 | 0.00842 |
| East Europe | Russian Federation (RU) | 1365 | 170856 | 0.00799 |
| East Europe | Lithuania (LT) | 375 | 47921 | 0.00783 |
| East Europe | Moldova (MD) | 53 | 7888 | 0.00672 |
| East Europe | Ukraine (UA) | 840 | 125052 | 0.00672 |
| East Europe | Israel (IL) | 66 | 10496 | 0.00629 |
| East Europe | Poland (PL) | 1202 | 241010 | 0.00499 |
| East Europe | Austria (AT) | 214 | 47451 | 0.00451 |
| East Europe | Romania (RO) | 205 | 55838 | 0.00367 |
| East Europe | Hungary (HU) | 208 | 65488 | 0.00318 |
| East Europe | Tunisia (TN) | 11 | 4717 | 0.00233 |
| East Europe | Morocco (MA) | 29 | 13286 | 0.00218 |
| East Europe | South Africa (ZA) | 45 | 26238 | 0.00172 |
| East Europe | Argentina (AR) | 37 | 27224 | 0.00136 |
| East Europe | Czechia (CZ) | 44 | 37582 | 0.00117 |
| East Europe | Iraq (IQ) | 13 | 13319 | 0.00098 |
| East Europe | Germany (DE) | 265 | 282663 | 0.00094 |
| East Europe | Syrian Arab Republic (SY) | 15 | 16238 | 0.00092 |
| East Europe | Slovakia (SK) | 23 | 26858 | 0.00086 |
| East Europe | Turkey (TR) | 37 | 45426 | 0.00081 |
| East Europe | Australia (AU) | 31 | 44830 | 0.00069 |
| East Europe | Egypt (EG) | 11 | 18405 | 0.0006 |
| East Europe | Greece (GR) | 40 | 69499 | 0.00058 |
| East Europe | Canada (CA) | 94 | 215749 | 0.00044 |
| East Europe | United States of America (US) | 3775 | 8681625 | 0.00043 |
| East Europe | Switzerland (CH) | 11 | 29143 | 0.00038 |
| East Europe | Netherlands (NL) | 17 | 46768 | 0.00036 |
| East Europe | Great Britain (GB) | 237 | 666326 | 0.00036 |
| East Europe | Norway (NO) | 18 | 58792 | 0.00031 |
| East Europe | France (FR) | 26 | 88880 | 0.00029 |
| East Europe | Italy (IT) | 108 | 425699 | 0.00025 |
| East Europe | Ireland (IE) | 47 | 202512 | 0.00023 |
| East Europe | Spain (ES) | 21 | 98925 | 0.00021 |

| | | | | |
|---|---|---|---|---|
| East Europe | Brazil (BR) | 14 | 70276 | 0.0002 |
| East Europe | Sweden (SE) | 10 | 64080 | 0.00016 |
| East Europe | Cuba (CU) | 10 | 95369 | 0.0001 |
| East Europe | Puerto Rico (PR) | 17 | 172278 | 0.0001 |
| East Europe | Mexico (MX) | 12 | 510699 | 0.00002 |
| Italy | Italy (IT) | 213 | 425699 | 0.0005 |
| Italy | United States of America (US) | 144 | 8681625 | 0.00002 |
| Mexico | Mexico (MX) | 613 | 510699 | 0.0012 |
| Mexico | Spain (ES) | 30 | 98925 | 0.0003 |
| Mexico | United States of America (US) | 165 | 8681625 | 0.00002 |
| North Africa | Algeria (DZ) | 136 | 8243 | 0.0165 |
| North Africa | Morocco (MA) | 209 | 13286 | 0.01573 |
| North Africa | Libya (LY) | 21 | 2033 | 0.01033 |
| North Africa | Tunisia (TN) | 45 | 4717 | 0.00954 |
| North Africa | Syrian Arab Republic (SY) | 20 | 16238 | 0.00123 |
| North Africa | Greece (GR) | 12 | 69499 | 0.00017 |
| North Africa | Italy (IT) | 12 | 425699 | 0.00003 |
| North Africa | Great Britain(GB) | 10 | 666326 | 0.00002 |
| North Africa | United States of America (US) | 26 | 8681625 | 0 |
| Northwestern Europe | Norway (NO) | 20 | 58792 | 0.00034 |
| Northwestern Europe | Sweden (SE) | 16 | 64080 | 0.00025 |
| Northwestern Europe | Great Britain(GB) | 153 | 666326 | 0.00023 |
| Northwestern Europe | United States of America (US) | 1807 | 8681625 | 0.00021 |
| Northwestern Europe | Italy (IT) | 78 | 425699 | 0.00018 |
| Northwestern Europe | Ireland (IE) | 37 | 202512 | 0.00018 |
| Northwestern Europe | Germany (DE) | 42 | 282663 | 0.00015 |
| Northwestern Europe | Canada (CA) | 31 | 215749 | 0.00014 |
| Northwestern Europe | Spain (ES) | 14 | 98925 | 0.00014 |
| Northwestern Europe | Brazil (BR) | 10 | 70276 | 0.00014 |
| Puerto Rico | Puerto Rico (PR) | 799 | 172278 | 0.00464 |
| Puerto Rico | Dominican Republic (DO) | 18 | 56217 | 0.00032 |
| Puerto Rico | Cuba (CU) | 26 | 95369 | 0.00027 |
| Puerto Rico | Portugal (PT) | 14 | 63740 | 0.00022 |
| Puerto Rico | Spain (ES) | 21 | 98925 | 0.00021 |
| Puerto Rico | Mexico (MX) | 14 | 510699 | 0.00003 |
| Puerto Rico | Italy (IT) | 13 | 425699 | 0.00003 |
| Puerto Rico | United States of America (US) | 194 | 8681625 | 0.00002 |

*Note*. Carrier frequency is the number of grandparents reported to have been born in a country by LRRK2 G2019S carriers (G2019S carrier count) divided by the total number of grandparents reported to have been born in a country by the entire 23andMe participant database (total participant count). To protect participant privacy, we excluded countries with small counts ($n$<5).

Supplementary Table 4. *Symptom Prevalence Between LRRK2 G2019S PD and Non-carrier PD Adjusting for Baseline Age, Disease Duration, and Education.*

| | | | | 95% CI | | | | |
|---|---|---|---|---|---|---|---|---|
| **Domain** | **Symptom** | **Term** | ***OR*** | ***LL*** | ***UL*** | ***SE*** | ***Z*** | $p_{FDR}$ |
| Autonomic | Constipation | Intercept | 0.16 | 0.04 | 0.55 | 0.64 | -2.87 | 0.017 |
| | | Symptom | 0.70 | 0.40 | 1.16 | 0.27 | -1.30 | 0.271 |
| | | Baseline Age | 0.98 | 0.97 | 1.00 | 0.01 | -1.79 | 0.144 |
| | | Education | 1.71 | 1.16 | 2.61 | 0.21 | 2.63 | 0.037 |
| | | Disease Duration | 1.04 | 1.01 | 1.06 | 0.01 | 2.62 | 0.011 |
| | Erectile Dysfunction | Intercept | 0.06 | 0.01 | 0.41 | 1.02 | -2.78 | 0.017 |
| | | Symptom | 0.76 | 0.48 | 1.21 | 0.24 | -1.14 | 0.334 |
| | | Baseline Age | 1.00 | 0.97 | 1.03 | 0.01 | -0.13 | 0.898 |
| | | Education | 1.33 | 0.74 | 2.58 | 0.32 | 0.90 | 0.369 |
| | | Disease Duration | 1.05 | 1.01 | 1.09 | 0.02 | 2.56 | 0.012 |
| | Nocturia | Intercept | 0.16 | 0.04 | 0.54 | 0.64 | -2.88 | 0.017 |
| | | Symptom | 1.13 | 0.75 | 1.65 | 0.20 | 0.59 | 0.582 |
| | | Baseline Age | 0.98 | 0.97 | 1.00 | 0.01 | -1.81 | 0.144 |
| | | Education | 1.62 | 1.10 | 2.45 | 0.20 | 2.37 | 0.037 |
| | | Disease Duration | 1.04 | 1.01 | 1.06 | 0.01 | 2.78 | 0.007 |
| | Orthostatic Hypotension | Intercept | 0.17 | 0.05 | 0.63 | 0.66 | -2.64 | 0.017 |
| | | Symptom | 0.73 | 0.52 | 1.00 | 0.16 | -1.97 | 0.073 |
| | | Baseline Age | 0.98 | 0.97 | 1.00 | 0.01 | -1.70 | 0.144 |
| | | Education | 1.57 | 1.07 | 2.38 | 0.20 | 2.23 | 0.039 |
| | | Disease Duration | 1.04 | 1.01 | 1.07 | 0.01 | 2.98 | 0.007 |
| | Urinary Difficulties | Intercept | 0.13 | 0.03 | 0.46 | 0.66 | -3.10 | 0.017 |
| | | Symptom | 0.88 | 0.64 | 1.20 | 0.16 | -0.82 | 0.458 |
| | | Baseline Age | 0.99 | 0.97 | 1.01 | 0.01 | -1.38 | 0.175 |
| | | Education | 1.67 | 1.13 | 2.54 | 0.21 | 2.49 | 0.037 |
| | | Disease Duration | 1.04 | 1.01 | 1.07 | 0.01 | 3.02 | 0.007 |
| Cognition | Attention/ Concentrating | Intercept | 0.26 | 0.07 | 0.96 | 0.67 | -1.99 | 0.048 |
| | | Symptom | 0.50 | 0.36 | 0.70 | 0.17 | -3.97 | 2.02E-04 |
| | | Baseline Age | 0.98 | 0.96 | 1.00 | 0.01 | -2.11 | 0.144 |
| | | Education | 1.55 | 1.04 | 2.36 | 0.21 | 2.11 | 0.042 |
| | | Disease Duration | 1.04 | 1.02 | 1.07 | 0.01 | 3.11 | 0.007 |
| | Distracted Easily | Intercept | 0.27 | 0.07 | 1.02 | 0.68 | -1.90 | 0.057 |
| | | Symptom | 0.51 | 0.36 | 0.71 | 0.17 | -3.86 | 2.60E-04 |
| | | Baseline Age | 0.98 | 0.96 | 1.00 | 0.01 | -2.17 | 0.144 |
| | | Education | 1.64 | 1.10 | 2.54 | 0.21 | 2.33 | 0.037 |

| | | | | | | | | |
|---|---|---|---|---|---|---|---|---|
| | | Disease Duration | 1.04 | 1.01 | 1.07 | 0.01 | 2.92 | 0.007 |
| | MCI | Intercept | 0.18 | 0.05 | 0.64 | 0.66 | -2.60 | 0.018 |
| | | Symptom | 0.28 | 0.10 | 0.62 | 0.46 | -2.77 | 0.011 |
| | | Baseline Age | 0.98 | 0.97 | 1.00 | 0.01 | -1.75 | 0.144 |
| | | Education | 1.48 | 1.00 | 2.24 | 0.20 | 1.91 | 0.059 |
| | | Disease Duration | 1.04 | 1.01 | 1.07 | 0.01 | 2.98 | 0.007 |
| | Multitasking | Intercept | 0.25 | 0.07 | 0.91 | 0.67 | -2.07 | 0.042 |
| | | Symptom | 0.41 | 0.30 | 0.57 | 0.17 | -5.23 | 3.65E-06 |
| | | Baseline Age | 0.98 | 0.97 | 1.00 | 0.01 | -1.73 | 0.144 |
| | | Education | 1.52 | 1.02 | 2.33 | 0.21 | 1.97 | 0.054 |
| | | Disease Duration | 1.04 | 1.01 | 1.07 | 0.01 | 2.85 | 0.007 |
| | Task Completion | Intercept | 0.23 | 0.06 | 0.83 | 0.67 | -2.22 | 0.031 |
| | | Symptom | 0.43 | 0.28 | 0.64 | 0.21 | -4.00 | 2.02E-04 |
| | | Baseline Age | 0.98 | 0.96 | 1.00 | 0.01 | -2.08 | 0.144 |
| | | Education | 1.72 | 1.15 | 2.67 | 0.22 | 2.52 | 0.037 |
| | | Disease Duration | 1.04 | 1.01 | 1.07 | 0.01 | 2.87 | 0.007 |
| | Understanding | Intercept | 0.16 | 0.04 | 0.59 | 0.67 | -2.72 | 0.017 |
| | | Symptom | 0.40 | 0.26 | 0.60 | 0.21 | -4.29 | 7.47E-05 |
| | | Baseline Age | 0.99 | 0.97 | 1.01 | 0.01 | -1.39 | 0.175 |
| | | Education | 1.55 | 1.04 | 2.38 | 0.21 | 2.10 | 0.042 |
| | | Disease Duration | 1.04 | 1.01 | 1.07 | 0.01 | 2.87 | 0.007 |
| Memory | Finding the Right Word | Intercept | 0.17 | 0.05 | 0.63 | 0.66 | -2.64 | 0.017 |
| | | Symptom | 0.67 | 0.48 | 0.93 | 0.17 | -2.44 | 0.026 |
| | | Baseline Age | 0.99 | 0.97 | 1.00 | 0.01 | -1.49 | 0.175 |
| | | Education | 1.68 | 1.14 | 2.58 | 0.21 | 2.50 | 0.037 |
| | | Disease Duration | 1.03 | 1.01 | 1.06 | 0.01 | 2.52 | 0.013 |
| | Forgetting the Date | Intercept | 0.17 | 0.05 | 0.61 | 0.66 | -2.68 | 0.017 |
| | | Symptom | 0.41 | 0.27 | 0.60 | 0.20 | -4.45 | 5.00E-05 |
| | | Baseline Age | 0.99 | 0.97 | 1.00 | 0.01 | -1.59 | 0.157 |
| | | Education | 1.56 | 1.06 | 2.39 | 0.21 | 2.16 | 0.040 |
| | | Disease Duration | 1.05 | 1.02 | 1.07 | 0.01 | 3.40 | 0.005 |
| | Misplacing Things | Intercept | 0.20 | 0.05 | 0.76 | 0.69 | -2.34 | 0.025 |
| | | Symptom | 0.46 | 0.31 | 0.67 | 0.20 | -3.95 | 2.02E-04 |
| | | Baseline Age | 0.98 | 0.96 | 1.00 | 0.01 | -1.88 | 0.144 |
| | | Education | 1.72 | 1.14 | 2.67 | 0.22 | 2.50 | 0.037 |
| | | Disease Duration | 1.05 | 1.02 | 1.07 | 0.01 | 3.24 | 0.006 |
| | Worsened Memory | Intercept | 0.21 | 0.05 | 0.81 | 0.70 | -2.23 | 0.031 |
| | | Symptom | 0.56 | 0.40 | 0.79 | 0.17 | -3.38 | 0.002 |
| | | Baseline Age | 0.98 | 0.96 | 1.00 | 0.01 | -1.70 | 0.144 |
| | | Education | 1.70 | 1.12 | 2.70 | 0.22 | 2.38 | 0.037 |

| | | | | | | | | |
|---|---|---|---|---|---|---|---|---|
| | | Disease Duration | 1.04 | 1.01 | 1.07 | 0.01 | 2.51 | 0.013 |
| Motor | Bradykinesia | Intercept | 0.19 | 0.05 | 0.67 | 0.65 | -2.55 | 0.019 |
| | | Symptom | 0.70 | 0.51 | 0.97 | 0.16 | -2.20 | 0.045 |
| | | Baseline Age | 0.98 | 0.97 | 1.00 | 0.01 | -1.74 | 0.144 |
| | | Education | 1.60 | 1.09 | 2.42 | 0.20 | 2.30 | 0.037 |
| | | Disease Duration | 1.04 | 1.01 | 1.07 | 0.01 | 2.91 | 0.007 |
| | Postural Instability | Intercept | 0.12 | 0.03 | 0.44 | 0.66 | -3.18 | 0.017 |
| | | Symptom | 0.96 | 0.69 | 1.36 | 0.17 | -0.22 | 0.823 |
| | | Baseline Age | 0.99 | 0.97 | 1.01 | 0.01 | -1.39 | 0.175 |
| | | Education | 1.64 | 1.11 | 2.50 | 0.21 | 2.40 | 0.037 |
| | | Disease Duration | 1.04 | 1.01 | 1.07 | 0.01 | 2.92 | 0.007 |
| | Reduced Arm Swing | Intercept | 0.17 | 0.04 | 0.68 | 0.73 | -2.45 | 0.020 |
| | | Symptom | 0.80 | 0.53 | 1.23 | 0.21 | -1.06 | 0.351 |
| | | Baseline Age | 0.99 | 0.97 | 1.01 | 0.01 | -1.42 | 0.175 |
| | | Education | 1.71 | 1.13 | 2.68 | 0.22 | 2.44 | 0.037 |
| | | Disease Duration | 1.03 | 1.00 | 1.06 | 0.01 | 2.16 | 0.031 |
| | Tremor | Intercept | 0.18 | 0.05 | 0.68 | 0.68 | -2.49 | 0.019 |
| | | Symptom | 0.82 | 0.56 | 1.21 | 0.20 | -1.04 | 0.351 |
| | | Baseline Age | 0.98 | 0.97 | 1.00 | 0.01 | -1.73 | 0.144 |
| | | Education | 1.56 | 1.06 | 2.36 | 0.20 | 2.19 | 0.040 |
| | | Disease Duration | 1.04 | 1.01 | 1.06 | 0.01 | 2.77 | 0.007 |
| RBD | RBD | Intercept | 0.16 | 0.04 | 0.57 | 0.65 | -2.81 | 0.017 |
| | | Symptom | 0.31 | 0.18 | 0.51 | 0.26 | -4.43 | 5.00E-05 |
| | | Baseline Age | 0.99 | 0.97 | 1.00 | 0.01 | -1.63 | 0.154 |
| | | Education | 1.58 | 1.07 | 2.40 | 0.21 | 2.23 | 0.039 |
| | | Disease Duration | 1.05 | 1.03 | 1.08 | 0.01 | 3.89 | 0.002 |
| Smell | Olfactory Difficulties | Intercept | 0.20 | 0.06 | 0.69 | 0.64 | -2.51 | 0.019 |
| | | Symptom | 0.43 | 0.31 | 0.59 | 0.17 | -5.08 | 3.86E-06 |
| | | Baseline Age | 0.98 | 0.97 | 1.00 | 0.01 | -1.82 | 0.144 |
| | | Education | 1.76 | 1.19 | 2.66 | 0.20 | 2.76 | 0.037 |
| | | Disease Duration | 1.05 | 1.02 | 1.07 | 0.01 | 3.45 | 0.005 |

*Note*. MCI=mild cognitive impairment; RBD=REM sleep behavior disorder; OR=odds ratio; CI = confidence interval; LL=lower limit; UL=upper limit; SE=Standard error; Z=Z value, $p_{FDR}$=p-values adjusted for false discovery rate.

Supplementary Table 5. *Logistic Regression Coefficients and Relative Odds of PD Across LRRK2 G2019S Carriers and Non-carriers: Comparing PRS With and Without the LRRK2 Locus in All Participants.*

| | | | | 95% CI | | | | |
|---|---|---|---|---|---|---|---|---|
| **PRS** | **Carrier Status** | **Term** | ***OR*** | ***LL*** | ***UL*** | ***SE*** | ***Z*** | ***p*** |
| Original | Non-carrier | Intercept | 0.01 | 0.01 | 0.02 | 0.037 | -115.43 | 0.00E+00 |
| | | PRS | 1.63 | 1.56 | 1.70 | 0.022 | 21.82 | 1.47E-105 |
| | | Age | 1.93 | 1.84 | 2.02 | 0.025 | 26.76 | 1.01E-157 |
| | | Male | 1.78 | 1.63 | 1.94 | 0.045 | 12.92 | 3.62E-38 |
| | | PC1 | 0.92 | 0.86 | 0.98 | 0.032 | -2.61 | 0.009 |
| | | PC2 | 0.94 | 0.88 | 1.00 | 0.033 | -1.86 | 0.062 |
| | | PC3 | 1.05 | 0.99 | 1.12 | 0.033 | 1.50 | 0.133 |
| | | PC4 | 0.97 | 0.93 | 1.02 | 0.025 | -1.13 | 0.259 |
| | | PC5 | 1.01 | 0.96 | 1.08 | 0.029 | 0.37 | 0.712 |
| | | PC6 | 1.11 | 1.05 | 1.17 | 0.028 | 3.62 | 2.91E-04 |
| | | PC7 | 0.99 | 0.93 | 1.06 | 0.033 | -0.18 | 0.855 |
| | | PC8 | 1.05 | 1.01 | 1.10 | 0.022 | 2.40 | 0.017 |
| | | PC9 | 0.97 | 0.92 | 1.02 | 0.028 | -1.19 | 0.235 |
| | | PC10 | 0.96 | 0.92 | 1.01 | 0.025 | -1.45 | 0.146 |
| | LRRK2 G2019S | Intercept | 0.19 | 0.15 | 0.24 | 0.127 | -13.22 | 7.11E-40 |
| | | PRS | 1.65 | 1.37 | 1.99 | 0.094 | 5.29 | 1.21E-07 |
| | | Age | 2.44 | 1.99 | 3.04 | 0.108 | 8.28 | 1.23E-16 |
| | | Male | 0.99 | 0.69 | 1.40 | 0.180 | -0.08 | 0.935 |
| | | PC1 | 1.06 | 0.72 | 1.50 | 0.182 | 0.29 | 0.769 |
| | | PC2 | 0.92 | 0.66 | 1.25 | 0.160 | -0.55 | 0.585 |
| | | PC3 | 1.07 | 0.75 | 1.56 | 0.182 | 0.40 | 0.692 |
| | | PC4 | 0.67 | 0.40 | 1.11 | 0.260 | -1.55 | 0.122 |
| | | PC5 | 1.41 | 0.85 | 2.36 | 0.260 | 1.33 | 0.183 |
| | | PC6 | 1.19 | 0.78 | 1.84 | 0.220 | 0.80 | 0.421 |
| | | PC7 | 0.93 | 0.58 | 1.48 | 0.238 | -0.29 | 0.775 |
| | | PC8 | 0.93 | 0.62 | 1.41 | 0.209 | -0.34 | 0.732 |
| | | PC9 | 1.11 | 0.92 | 1.33 | 0.093 | 1.10 | 0.272 |
| | | PC10 | 0.84 | 0.66 | 1.08 | 0.126 | -1.35 | 0.178 |
| Modified | Non-carrier | Intercept | 0.01 | 0.01 | 0.02 | 0.037 | -115.46 | 0.00E+00 |
| | | PRS | 1.62 | 1.55 | 1.69 | 0.022 | 21.62 | 1.26E-103 |
| | | Age | 1.93 | 1.84 | 2.02 | 0.025 | 26.75 | 1.20E-157 |
| | | Male | 1.78 | 1.63 | 1.94 | 0.045 | 12.88 | 5.58E-38 |
| | | PC1 | 0.92 | 0.86 | 0.98 | 0.032 | -2.53 | 0.012 |
| | | PC2 | 0.94 | 0.88 | 1.00 | 0.033 | -1.80 | 0.072 |
| | | PC3 | 1.04 | 0.98 | 1.12 | 0.033 | 1.33 | 0.184 |
| | | PC4 | 0.98 | 0.93 | 1.03 | 0.025 | -0.86 | 0.388 |
| | | PC5 | 1.01 | 0.96 | 1.07 | 0.029 | 0.32 | 0.752 |
| | | PC6 | 1.11 | 1.05 | 1.17 | 0.028 | 3.59 | 3.28E-04 |
| | | PC7 | 1.00 | 0.93 | 1.06 | 0.032 | -0.14 | 0.886 |
| | | PC8 | 1.05 | 1.01 | 1.10 | 0.022 | 2.35 | 0.019 |
| | | PC9 | 0.97 | 0.92 | 1.02 | 0.028 | -1.16 | 0.244 |
| | | PC10 | 0.96 | 0.92 | 1.01 | 0.025 | -1.51 | 0.132 |

| | | | | | | | |
|---|---|---|---|---|---|---|---|
| LRRK2 G2019S | Intercept | 0.19 | 0.15 | 0.24 | 0.126 | -13.22 | 7.10E-40 |
| | PRS | 1.56 | 1.31 | 1.87 | 0.090 | 4.97 | 6.59E-07 |
| | Age | 2.44 | 1.99 | 3.04 | 0.108 | 8.27 | 1.35E-16 |
| | Male | 0.97 | 0.68 | 1.38 | 0.179 | -0.15 | 0.884 |
| | PC1 | 1.05 | 0.71 | 1.48 | 0.180 | 0.25 | 0.801 |
| | PC2 | 0.91 | 0.65 | 1.24 | 0.160 | -0.61 | 0.541 |
| | PC3 | 1.07 | 0.74 | 1.54 | 0.181 | 0.35 | 0.725 |
| | PC4 | 0.68 | 0.40 | 1.12 | 0.260 | -1.51 | 0.132 |
| | PC5 | 1.42 | 0.86 | 2.38 | 0.259 | 1.36 | 0.174 |
| | PC6 | 1.23 | 0.80 | 1.89 | 0.219 | 0.94 | 0.345 |
| | PC7 | 0.93 | 0.58 | 1.47 | 0.237 | -0.30 | 0.765 |
| | PC8 | 0.95 | 0.63 | 1.44 | 0.208 | -0.25 | 0.799 |
| | PC9 | 1.10 | 0.92 | 1.32 | 0.093 | 1.01 | 0.312 |
| | PC10 | 0.85 | 0.66 | 1.09 | 0.125 | -1.28 | 0.199 |

*Note.* Age and PCs were Z-scored prior to modeling and are interpreted in *SD* units. PRS=polygenic risk score; OR=odds ratio; CI=confidence interval; LL=lower level; UL=upper level; SE=standard error; Z=Z-value; PC=principal component.

Supplementary Table 6. *Logistic Regression Coefficients and Relative Odds of PD Across LRRK2 G2019S Carriers and Non-carriers: Comparing PRS With and Without the LRRK2 Locus in Participants with European Ancestry.*

| | | | | **95% CI** | | | | |
|---|---|---|---|---|---|---|---|---|
| **PRS** | **Carrier Status** | **Term** | ***OR*** | ***LL*** | ***UL*** | ***SE*** | ***Z*** | ***p*** |
| Original | Non-carrier | Intercept | 0.02 | 0.01 | 0.02 | 0.038 | -109.54 | 0.00E+00 |
| | | PRS | 1.63 | 1.56 | 1.71 | 0.023 | 21.00 | 6.74E-98 |
| | | Age | 1.88 | 1.79 | 1.97 | 0.026 | 24.69 | 1.28E-134 |
| | | Male | 1.80 | 1.64 | 1.97 | 0.047 | 12.59 | 2.33E-36 |
| | | PC1 | 0.99 | 0.95 | 1.04 | 0.021 | -0.25 | 0.802 |
| | | PC2 | 0.94 | 0.90 | 0.98 | 0.022 | -3.01 | 0.003 |
| | | PC3 | 1.08 | 1.03 | 1.13 | 0.024 | 3.10 | 0.002 |
| | | PC4 | 1.01 | 0.96 | 1.05 | 0.022 | 0.33 | 0.741 |
| | | PC5 | 0.98 | 0.94 | 1.03 | 0.023 | -0.81 | 0.419 |
| | LRRK2 G2019S | Intercept | 0.21 | 0.16 | 0.27 | 0.134 | -11.76 | 6.21E-32 |
| | | PRS | 1.76 | 1.44 | 2.16 | 0.103 | 5.49 | 3.95E-08 |
| | | Age | 2.54 | 2.04 | 3.22 | 0.117 | 7.99 | 1.31E-15 |
| | | Male | 0.95 | 0.65 | 1.38 | 0.191 | -0.28 | 0.780 |
| | | PC1 | 0.85 | 0.61 | 1.17 | 0.165 | -0.98 | 0.327 |
| | | PC2 | 0.94 | 0.78 | 1.15 | 0.100 | -0.60 | 0.549 |
| | | PC3 | 1.10 | 0.80 | 1.57 | 0.170 | 0.58 | 0.560 |
| | | PC4 | 1.06 | 0.88 | 1.29 | 0.098 | 0.63 | 0.530 |
| | | PC5 | 1.09 | 0.90 | 1.32 | 0.098 | 0.85 | 0.396 |
| Modified | Non-carrier | Intercept | 0.02 | 0.01 | 0.02 | 0.038 | -109.56 | 0.00E+00 |
| | | PRS | 1.63 | 1.56 | 1.71 | 0.023 | 20.83 | 2.10E-96 |
| | | Age | 1.88 | 1.79 | 1.97 | 0.026 | 24.69 | 1.39E-134 |
| | | Male | 1.79 | 1.64 | 1.97 | 0.047 | 12.56 | 3.33E-36 |
| | | PC1 | 1.00 | 0.96 | 1.04 | 0.021 | -0.01 | 0.989 |
| | | PC2 | 0.94 | 0.90 | 0.98 | 0.022 | -3.01 | 0.003 |
| | | PC3 | 1.08 | 1.03 | 1.13 | 0.024 | 3.06 | 0.002 |
| | | PC4 | 1.01 | 0.96 | 1.05 | 0.022 | 0.31 | 0.758 |
| | | PC5 | 0.98 | 0.94 | 1.02 | 0.023 | -0.84 | 0.400 |
| | LRRK2 G2019S | Intercept | 0.21 | 0.16 | 0.27 | 0.133 | -11.72 | 1.04E-31 |
| | | PRS | 1.65 | 1.36 | 2.00 | 0.098 | 5.10 | 3.39E-07 |
| | | Age | 2.53 | 2.03 | 3.21 | 0.117 | 7.98 | 1.45E-15 |
| | | Male | 0.93 | 0.64 | 1.35 | 0.190 | -0.39 | 0.698 |
| | | PC1 | 0.86 | 0.62 | 1.19 | 0.165 | -0.90 | 0.367 |
| | | PC2 | 0.93 | 0.77 | 1.14 | 0.100 | -0.75 | 0.452 |
| | | PC3 | 1.15 | 0.84 | 1.63 | 0.169 | 0.85 | 0.398 |
| | | PC4 | 1.07 | 0.88 | 1.30 | 0.098 | 0.67 | 0.500 |
| | | PC5 | 1.08 | 0.90 | 1.31 | 0.097 | 0.83 | 0.407 |

*Note.* Age and PCs were Z-scored prior to modeling and are interpreted in *SD* units. PRS=polygenic risk score; OR=odds ratio; CI=confidence interval; LL=lower level; UL=upper level; SE=standard error; Z=Z-value; PC=principal component.

Supplementary Table 7. *Logistic Regression Coefficients and Relative Odds of PD Across Low, Intermediate, and High Modified PRS between LRRK2 G2019S Carriers and Non-carriers in All Participants.*

| | | **95% CI** | | | | |
|---|---|---|---|---|---|---|
| **Term** | ***OR*** | ***LL*** | ***UL*** | ***SE*** | ***Z*** | ***p*** |
| Intercept | 0.01 | 0.01 | 0.02 | 0.043 | -99.92 | 0.00E+00 |
| Non-carrier Low | 0.63 | 0.55 | 0.72 | 0.067 | -6.93 | 4.08E-12 |
| Non-carrier Inter. | 1.00 | | | | | |
| Non-carrier High | 1.95 | 1.78 | 2.14 | 0.048 | 13.98 | 2.03E-44 |
| LRRK2 G2019S Low | 6.06 | 3.71 | 9.49 | 0.239 | 7.55 | 4.24E-14 |
| LRRK2 G2019S Inter. | 9.09 | 6.82 | 11.99 | 0.144 | 15.36 | 2.94E-53 |
| LRRK2 G2019S High | 16.87 | 12.55 | 22.55 | 0.149 | 18.92 | 7.73E-80 |
| Age | 1.95 | 1.86 | 2.05 | 0.024 | 28.04 | 5.76E-173 |
| Male | 1.70 | 1.56 | 1.85 | 0.043 | 12.36 | 4.23E-35 |
| PC1 | 0.92 | 0.87 | 0.98 | 0.031 | -2.50 | 0.012 |
| PC2 | 0.94 | 0.88 | 1.00 | 0.033 | -1.80 | 0.072 |
| PC3 | 1.05 | 0.98 | 1.12 | 0.032 | 1.38 | 0.167 |
| PC4 | 0.97 | 0.92 | 1.02 | 0.026 | -1.04 | 0.297 |
| PC5 | 1.01 | 0.96 | 1.08 | 0.030 | 0.45 | 0.654 |
| PC6 | 1.10 | 1.04 | 1.16 | 0.027 | 3.44 | 0.001 |
| PC7 | 0.99 | 0.93 | 1.06 | 0.032 | -0.19 | 0.847 |
| PC8 | 1.05 | 1.01 | 1.10 | 0.022 | 2.35 | 0.019 |
| PC9 | 0.97 | 0.92 | 1.02 | 0.027 | -1.05 | 0.296 |
| PC10 | 0.96 | 0.92 | 1.01 | 0.024 | -1.50 | 0.132 |

*Note*. Groups were determined by modified PRS percentile ranges: Low (1-25%), Intermediate (25-75%), and High (75-100%); non-carriers with Intermediate PRS served as the reference group; Age and PCs were Z-scored prior to modeling and are interpreted in *SD* units; PRS=polygenic risk score; Inter.=Intermediate; OR=odds ratio; SE=standard error; Z=Z-value; CI=confidence interval; LL=lower level; UL=upper level; Inter.=intermediate; PC=principal component.

Supplementary Table 8. *Logistic Regression Coefficients and Relative Odds of PD Across Low, Intermediate, and High Modified PRS between LRRK2 G2019S Carriers and Non-carriers in European Participants.*

| | | **95% CI** | | | | |
|---|---|---|---|---|---|---|
| **Term** | ***OR*** | ***LL*** | ***UL*** | ***SE*** | ***Z*** | ***p*** |
| Intercept | 0.02 | 0.01 | 0.02 | 0.044 | -94.41 | 0.00E+00 |
| Non-carrier Low | 0.64 | 0.56 | 0.74 | 0.070 | -6.29 | 3.10E-10 |
| Non-carrier Inter. | 1.00 | | | | | |
| Non-carrier High | 2.01 | 1.82 | 2.21 | 0.050 | 13.95 | 2.98E-44 |
| LRRK2 G2019S Low | 5.53 | 3.17 | 9.14 | 0.269 | 6.37 | 1.89E-10 |
| LRRK2 G2019S Inter. | 8.80 | 6.49 | 11.83 | 0.153 | 14.21 | 7.93E-46 |
| LRRK2 G2019S High | 17.17 | 12.55 | 23.35 | 0.158 | 17.96 | 3.99E-72 |
| Age | 1.91 | 1.82 | 2.00 | 0.025 | 25.97 | 1.11E-148 |
| Male | 1.71 | 1.57 | 1.87 | 0.045 | 11.99 | 4.05E-33 |
| PC1 | 0.99 | 0.95 | 1.04 | 0.021 | -0.24 | 0.812 |
| PC2 | 0.94 | 0.90 | 0.98 | 0.022 | -2.87 | 0.004 |
| PC3 | 1.08 | 1.03 | 1.13 | 0.024 | 3.14 | 0.002 |
| PC4 | 1.01 | 0.97 | 1.05 | 0.022 | 0.36 | 0.722 |
| PC5 | 0.99 | 0.94 | 1.03 | 0.022 | -0.68 | 0.498 |

*Note*. Groups were determined by modified PRS percentile ranges: Low (1-25%), Intermediate (25-75%), and High (75-100%); non-carriers with Intermediate PRS served as the reference group; Age and PCs were Z-scored prior to modeling and are interpreted in *SD* units; PRS=polygenic risk score; Inter.=Intermediate; OR=odds ratio; SE=standard error; Z=Z-value; CI=confidence interval; LL=lower level; UL=upper level; Inter.=intermediate; PC=principal component.

Supplementary Table 9. *Logistic Regression Coefficients and Relative Odds of PD Across Deciles of Polygenic Risk Score in All Participants.*

| Term | *OR* | 95% CI *LL* | *UL* | *SE* | *Z* | *p* |
|---|---|---|---|---|---|---|
| Intercept | 0.01 | 0.01 | 0.01 | 0.083 | -52.52 | 0.00E+00 |
| Non-carrier - 1 | 0.57 | 0.44 | 0.73 | 0.128 | -4.40 | 1.11E-05 |
| Non-carrier - 2 | 0.82 | 0.66 | 1.03 | 0.116 | -1.67 | 0.095 |
| Non-carrier - 3 | 0.76 | 0.60 | 0.95 | 0.119 | -2.35 | 0.019 |
| Non-carrier - 4 | 0.98 | 0.79 | 1.22 | 0.111 | -0.16 | 0.872 |
| Non-carrier - 5 | 1.00 | | | | | |
| Non-carrier - 6 | 1.10 | 0.89 | 1.36 | 0.109 | 0.89 | 0.375 |
| Non-carrier - 7 | 1.34 | 1.10 | 1.65 | 0.104 | 2.84 | 0.005 |
| Non-carrier - 8 | 1.53 | 1.25 | 1.86 | 0.101 | 4.18 | 2.98E-05 |
| Non-carrier - 9 | 1.84 | 1.52 | 2.23 | 0.098 | 6.19 | 6.04E-10 |
| Non-carrier - 10 | 2.92 | 2.45 | 3.51 | 0.092 | 11.62 | 3.16E-31 |
| LRRK2 G2019S - 1 | 4.90 | 1.96 | 10.54 | 0.423 | 3.76 | 1.71E-04 |
| LRRK2 G2019S - 2 | 8.71 | 4.04 | 17.06 | 0.364 | 5.95 | 2.61E-09 |
| LRRK2 G2019S - 3 | 8.93 | 4.38 | 16.92 | 0.342 | 6.40 | 1.59E-10 |
| LRRK2 G2019S - 4 | 6.49 | 2.89 | 13.09 | 0.381 | 4.91 | 9.17E-07 |
| LRRK2 G2019S - 5 | 11.38 | 6.38 | 19.45 | 0.283 | 8.59 | 8.40E-18 |
| LRRK2 G2019S - 6 | 13.45 | 7.82 | 22.35 | 0.267 | 9.74 | 2.05E-22 |
| LRRK2 G2019S - 7 | 8.38 | 4.45 | 14.87 | 0.306 | 6.95 | 3.73E-12 |
| LRRK2 G2019S - 8 | 10.98 | 6.34 | 18.30 | 0.269 | 8.90 | 5.58E-19 |
| LRRK2 G2019S - 9 | 18.71 | 11.58 | 29.71 | 0.240 | 12.22 | 2.53E-34 |
| LRRK2 G2019S - 10 | 24.69 | 15.97 | 37.80 | 0.219 | 14.61 | 2.33E-48 |
| Age | 1.95 | 1.86 | 2.05 | 0.024 | 27.99 | 2.22E-172 |
| Male | 1.71 | 1.57 | 1.86 | 0.043 | 12.46 | 1.22E-35 |
| PC1 | 0.93 | 0.87 | 0.98 | 0.031 | -2.39 | 0.017 |
| PC2 | 0.94 | 0.88 | 1.00 | 0.033 | -1.82 | 0.068 |
| PC3 | 1.04 | 0.98 | 1.11 | 0.032 | 1.31 | 0.192 |
| PC4 | 0.97 | 0.92 | 1.02 | 0.026 | -1.10 | 0.271 |
| PC5 | 1.01 | 0.96 | 1.08 | 0.030 | 0.45 | 0.651 |
| PC6 | 1.10 | 1.05 | 1.16 | 0.027 | 3.58 | 3.49E-04 |
| PC7 | 1.00 | 0.93 | 1.06 | 0.032 | -0.13 | 0.900 |
| PC8 | 1.05 | 1.00 | 1.09 | 0.022 | 2.12 | 0.034 |
| PC9 | 0.97 | 0.92 | 1.03 | 0.027 | -0.99 | 0.323 |
| PC10 | 0.96 | 0.92 | 1.01 | 0.024 | -1.67 | 0.096 |

*Note*. Numbers to the right of groups indicate decile; non-carriers at the 5th decile of PRS served as the reference group; Age and PCs were Z-scored prior to modeling and are interpreted in *SD* units; OR=odds ratio; CI=confidence interval; LL=lower level; UL=upper level; SE=standard error; Z=Z-value; PC=principal component.

Supplementary Table 10. *Logistic Regression Coefficients and Relative Odds of PD Across Deciles of Polygenic Risk Score in European Participants.*

| | | **95% CI** | | | | |
|---|---|---|---|---|---|---|
| **Term** | ***OR*** | ***LL*** | ***UL*** | ***SE*** | ***Z*** | ***p*** |
| Intercept | 0.01 | 0.01 | 0.02 | 0.086 | -49.57 | 0.00E+00 |
| Non-carrier - 1 | 0.55 | 0.42 | 0.72 | 0.135 | -4.39 | 1.12E-05 |
| Non-carrier - 2 | 0.83 | 0.66 | 1.05 | 0.120 | -1.53 | 0.126 |
| Non-carrier - 3 | 0.73 | 0.57 | 0.93 | 0.124 | -2.49 | 0.013 |
| Non-carrier - 4 | 0.98 | 0.78 | 1.23 | 0.115 | -0.18 | 0.861 |
| Non-carrier - 5 | 1.00 | | | | | |
| Non-carrier - 6 | 1.08 | 0.86 | 1.35 | 0.113 | 0.67 | 0.501 |
| Non-carrier - 7 | 1.27 | 1.03 | 1.58 | 0.109 | 2.21 | 0.027 |
| Non-carrier - 8 | 1.43 | 1.16 | 1.76 | 0.106 | 3.38 | 0.001 |
| Non-carrier - 9 | 1.88 | 1.54 | 2.30 | 0.101 | 6.21 | 5.21E-10 |
| Non-carrier - 10 | 2.95 | 2.45 | 3.57 | 0.096 | 11.32 | 1.05E-29 |
| LRRK2 G2019S - 1 | 4.10 | 1.38 | 9.84 | 0.492 | 2.87 | 0.004 |
| LRRK2 G2019S - 2 | 8.38 | 3.51 | 17.80 | 0.409 | 5.19 | 2.05E-07 |
| LRRK2 G2019S - 3 | 7.59 | 3.49 | 15.09 | 0.370 | 5.47 | 4.39E-08 |
| LRRK2 G2019S - 4 | 6.15 | 2.73 | 12.44 | 0.383 | 4.74 | 2.11E-06 |
| LRRK2 G2019S - 5 | 12.40 | 6.75 | 21.88 | 0.299 | 8.43 | 3.53E-17 |
| LRRK2 G2019S - 6 | 12.78 | 7.20 | 21.84 | 0.282 | 9.05 | 1.49E-19 |
| LRRK2 G2019S - 7 | 8.51 | 4.39 | 15.54 | 0.321 | 6.68 | 2.37E-11 |
| LRRK2 G2019S - 8 | 7.76 | 4.11 | 13.82 | 0.308 | 6.66 | 2.82E-11 |
| LRRK2 G2019S - 9 | 18.09 | 11.06 | 29.11 | 0.246 | 11.76 | 6.49E-32 |
| LRRK2 G2019S - 10 | 27.11 | 17.11 | 42.66 | 0.233 | 14.19 | 1.10E-45 |
| Age | 1.91 | 1.82 | 2.00 | 0.025 | 25.92 | 4.05E-148 |
| Male | 1.73 | 1.58 | 1.88 | 0.045 | 12.10 | 1.12E-33 |
| PC1 | 0.99 | 0.95 | 1.04 | 0.022 | -0.30 | 0.761 |
| PC2 | 0.93 | 0.90 | 0.98 | 0.022 | -3.12 | 0.002 |
| PC3 | 1.07 | 1.03 | 1.13 | 0.024 | 3.04 | 0.002 |
| PC4 | 1.01 | 0.97 | 1.05 | 0.022 | 0.46 | 0.646 |
| PC5 | 0.98 | 0.94 | 1.03 | 0.022 | -0.69 | 0.493 |

*Note.* Numbers to the right of groups indicate decile; non-carriers at the 5th decile of PRS served as the reference group; Age and PCs were Z-scored prior to modeling and are interpreted in *SD* units; OR=odds ratio; CI=confidence interval; LL=lower level; UL=upper level; SE=standard error; Z=Z-value; PC=principal component.

### References

Aguet, F., Barbeira, A. N., Bonazzola, R., Brown, A., Castel, S. E., Jo, B., Kasela, S., Kim-Hellmuth, S., Liang, Y., Oliva, M., Parsana, P. E., Flynn, E., Fresard, L., Gaamzon, E. R., Hamel, A. R., He, Y., Hormozdiari, F., Mohammadi, P., Muñoz-Aguirre, M., … Lappalainen, T. (2019). *The GTEx Consortium atlas of genetic regulatory effects across human tissues* (p. 787903). bioRxiv. https://doi.org/10.1101/787903

Auton, A., Abecasis, G. R., Altshuler, D. M., Durbin, R. M., Abecasis, G. R., Bentley, D. R., Chakravarti, A., Clark, A. G., Donnelly, P., Eichler, E. E., Flicek, P., Gabriel, S. B., Gibbs, R. A., Green, E. D., Hurles, M. E., Knoppers, B. M., Korbel, J. O., Lander, E. S., Lee, C., … National Eye Institute, N. (2015). A global reference for human genetic variation. *Nature*, *526*(7571), Article 7571. https://doi.org/10.1038/nature15393

Barton, A. R., Sherman, M. A., Mukamel, R. E., & Loh, P.-R. (2021). Whole-exome imputation within UK Biobank powers rare coding variant association and fine-mapping analyses. *Nature Genetics*, *53*(8), Article 8. https://doi.org/10.1038/s41588-021-00892-1

Bergström, A., McCarthy, S. A., Hui, R., Almarri, M. A., Ayub, Q., Danecek, P., Chen, Y., Felkel, S., Hallast, P., Kamm, J., Blanché, H., Deleuze, J.-F., Cann, H., Mallick, S., Reich, D., Sandhu, M. S., Skoglund, P., Scally, A., Xue, Y., … Tyler-Smith, C. (2020). Insights into human genetic variation and population history from 929 diverse genomes. *Science*, *367*(6484), eaay5012. https://doi.org/10.1126/science.aay5012

Browning, B. L., Zhou, Y., & Browning, S. R. (2018). A One-Penny Imputed Genome from Next-Generation Reference Panels. *The American Journal of Human Genetics*, *103*(3), 338–348. https://doi.org/10.1016/j.ajhg.2018.07.015

Durand, E. Y., Do, C. B., Mountain, J. L., & Macpherson, J. M. (2014). *Ancestry Composition: A Novel, Efficient Pipeline for Ancestry Deconvolution* (p. 010512). bioRxiv. https://doi.org/10.1101/010512

Li, H., Bloom, J. M., Farjoun, Y., Fleharty, M., Gauthier, L., Neale, B., & MacArthur, D. (2018). A synthetic-diploid benchmark for accurate variant-calling evaluation. *Nature Methods*, *15*(8), Article 8. https://doi.org/10.1038/s41592-018-0054-7

Mallick, S., Li, H., Lipson, M., Mathieson, I., Gymrek, M., Racimo, F., Zhao, M., Chennagiri, N., Nordenfelt, S., Tandon, A., Skoglund, P., Lazaridis, I., Sankararaman, S., Fu, Q., Rohland, N., Renaud, G., Erlich, Y., Willems, T., Gallo, C., … Reich, D. (2016). The Simons Genome Diversity Project: 300 genomes from 142 diverse populations. *Nature*, *538*(7624), Article 7624. https://doi.org/10.1038/nature18964
